# National incidence of coccidioidomycosis in the United States estimated through machine learning integration of epidemiological, veterinary, and socioecological data

**DOI:** 10.64898/2026.10.14.26365564

**Authors:** Lisa I. Couper, Simon K. Camponuri, Jane E. Sykes, Xiudi Li, Alan E. Hubbard, Brittany L. Morgan Bustamante, Thomas Williamson, Irene Ruberto, Gail Sondermeyer-Cooksey, Ellora N. Karmarkar, Alexander T. Yu, Alexandra K. Heaney, Jennifer R. Head, Justin V. Remais

## Abstract

**Key points:** *Question:* In the absence of nationwide reporting for coccidioidomycosis, can alternative data sources be integrated to enable estimates of monthly, county-level incidence across the US?

*Findings:* In this cross-sectional study of 3,108 counties, the model identified 60 counties in non-reporting states with predicted incidence ≥2 cases per 100,000 population (considered moderate to high incidence). In states with mandatory reporting outside of Arizona and California, the model’s predicted county-level incidence rates were, on average, 28% higher than those captured in national surveillance.

*Meaning:* Our predictions highlight the extent of case under-ascertainment for coccidioidomycosis relative to Arizona and California, both in states with mandatory reporting and in non-reporting states.

**Importance:** Coccidioidomycosis, also known as Valley fever, is a serious fungal disease caused by inhaling *Coccidioides* spores aerosolized from the soil. Despite rising public health concern, the distribution and incidence of coccidioidomycosis in the US remain poorly characterized as not all states are required to report cases, and case under-ascertainment is presumed to be high, particularly outside of the endemic region.

**Objective:** To estimate monthly, county-level coccidioidomycosis incidence across the US while accounting for widespread under-ascertainment.

**Design, Setting, and Participants:** We integrated multisource data, including human cases diagnosed in electronic health records, serological testing of dogs, and ecological and sociodemographic predictors, in a machine learning model estimating monthly, county-level coccidioidomycosis incidence. The model was trained on surveillance data from Arizona and California from 2013 to 2023, then applied to predict incidence in states across the contiguous US over the same time period.

**Main Outcomes and Measures:** Incidence of coccidioidomycosis at the monthly county level.

**Results:** The model predicted coccidioidomycosis incidence from 2013 to 2023 for 3,108 counties from all states in the contiguous US. The model identified 60 counties in non-reporting states where predicted incidence was ≥2 cases per 100,000 population (considered moderate to high incidence). In states with mandatory reporting outside of Arizona and California, the model predicted county-level incidence rates that were 28% higher (95% bootstrap interval, 0.1%–69.8%) than those captured in national surveillance, equating to a total of 608 potentially missed cases per year (95% bootstrap interval, 340–964).

**Conclusions and Relevance:** In this cross-sectional study, the model quantified case under-ascertainment for coccidioidomycosis relative to Arizona and California, and highlighted the potential for alternative data sources to fill in gaps in mandatory reporting. Estimating incidence while accounting for under-ascertainment and non-reporting may help efforts to improve public and physician awareness and to detect emergence in new regions.

## Introduction

Coccidioidomycosis (commonly known as Valley fever) is a serious, emerging fungal disease caused by inhaling spores of the soil-dwelling fungus *Coccidioides*. While the majority of infections are asymptomatic or self-limiting, severe life-threatening disease can occur, including in otherwise healthy individuals^1,2^. Early diagnosis and proper management of symptomatic individuals are therefore critical for preventing severe complications and improving patient outcomes, particularly as there are currently no vaccines approved for human use. However, delayed diagnosis and misdiagnosis (including inappropriate use of antibiotics) are common^3^ as coccidioidomycosis symptoms can resemble those of more common illnesses (e.g., bacterial or viral community-acquired pneumonia, influenza, COVID-19)^4,5^, and awareness of the disease among clinicians and patients is limited, particularly outside of the highly endemic region^6–9^.

In the US, over 90% of coccidioidomycosis cases are reported in Arizona and California^10^, as *Coccidioides* grows in arid, desert-like soils^11^. However, locally acquired infections have recently been documented outside the historically recognized endemic zone, including in eastern Washington state^12^ and Oregon^13^, and the endemic region is projected to expand under changing environmental conditions^14,15^. To date, the national distribution of coccidioidomycosis incidence remains incompletely understood, in part due to a lack of nationwide reporting requirements. The primary method for tracking overall national and statewide coccidioidomycosis incidence and expansion in the US is public health reporting, which entails state-mandated reporting of newly diagnosed cases by health care providers or laboratories to local health departments. Yet nearly half of US states (n = 22)—including several where *Coccidioides* may be endemic, such as Idaho, Oklahoma, and Texas^6,16,17^—do not have such mandatory reporting^18^.

Cases may also go unreported due to barriers to accessing health care, misdiagnosis, and underreporting even when mandatory. A prior study estimated that only 60% of coccidioidomycosis patients seek medical care (including 30% of those with symptomatic, uncomplicated disease and 99% of those with severe disease)^6^. Moreover, among states with mandatory reporting outside of Arizona and California, fewer than 20% of patients with uncomplicated disease are estimated to be accurately diagnosed, and fewer than 32% of accurately diagnosed cases are estimated to be reported to public health entities^6^. Thus, even among states where coccidioidomycosis is reportable, national surveillance is estimated to capture less than 5% of cases outside of Arizona and California^6^.

Data sources besides state surveillance may help fill in gaps in coccidioidomycosis reporting and case ascertainment. For example, results from antigen-based skin test surveys conducted in the 1940s and 1950s^19^ formed the initial assessment of the coccidioidomycosis distribution in the US, which was then refined through case data from state public health departments, case investigations incorporating patient travel histories^13,20^, and/or genomic epidemiology^12,21–23^.

More recently, cases diagnosed in electronic health records (EHR) have been used to characterize trends in coccidioidomycosis^24^ as well as other fungal diseases in the US.^25,26^ Additionally, a nationwide dataset on serological testing for anti-*Coccidioides* antibodies in domestic dogs (*Canis familiaris*)—which have been proposed as sentinels for coccidioidomycosis^27^—was recently compiled, providing insight into the potential distribution of human disease^17^. Less directly, several prior studies have used climate and other ecological data to estimate the potential geographic distribution of *Coccidioides* in the US.^28–31^ While these alternative data sources have helped shed light on spatial and temporal characteristics of coccidioidomycosis, a systematic assessment of current coccidioidomycosis incidence across the US has remained out of reach as no single contemporary data stream provides national coverage while accounting for widespread under-ascertainment.

Here, we estimate monthly, county-level incidence of coccidioidomycosis in the US using a machine learning data integration approach. We combine data on human cases diagnosed in EHR, serological test results in dogs, and the early skin test reactivity results, alongside finely resolved ecological predictors of *Coccidioides* habitat suitability (e.g., monthly temperature and precipitation, drought severity, land cover, soil properties, dust concentrations), and sociodemographic predictors of exposure, disease severity, and human health care-seeking behavior (e.g., health insurance coverage, household income, percent employment in construction and extraction) (Table 1). We trained a model using these data sources to predict known census tract-level incident cases in Arizona and California, then applied the model to predict county-level cases across all states in the contiguous US. Our predictions thus represent the coccidioidomycosis incidence that would have been observed if all states had rates of health care seeking, accurate diagnosis, and case reporting akin to those of Arizona and California, which are estimated to be the highest in the nation^6^.

**Table 1.**
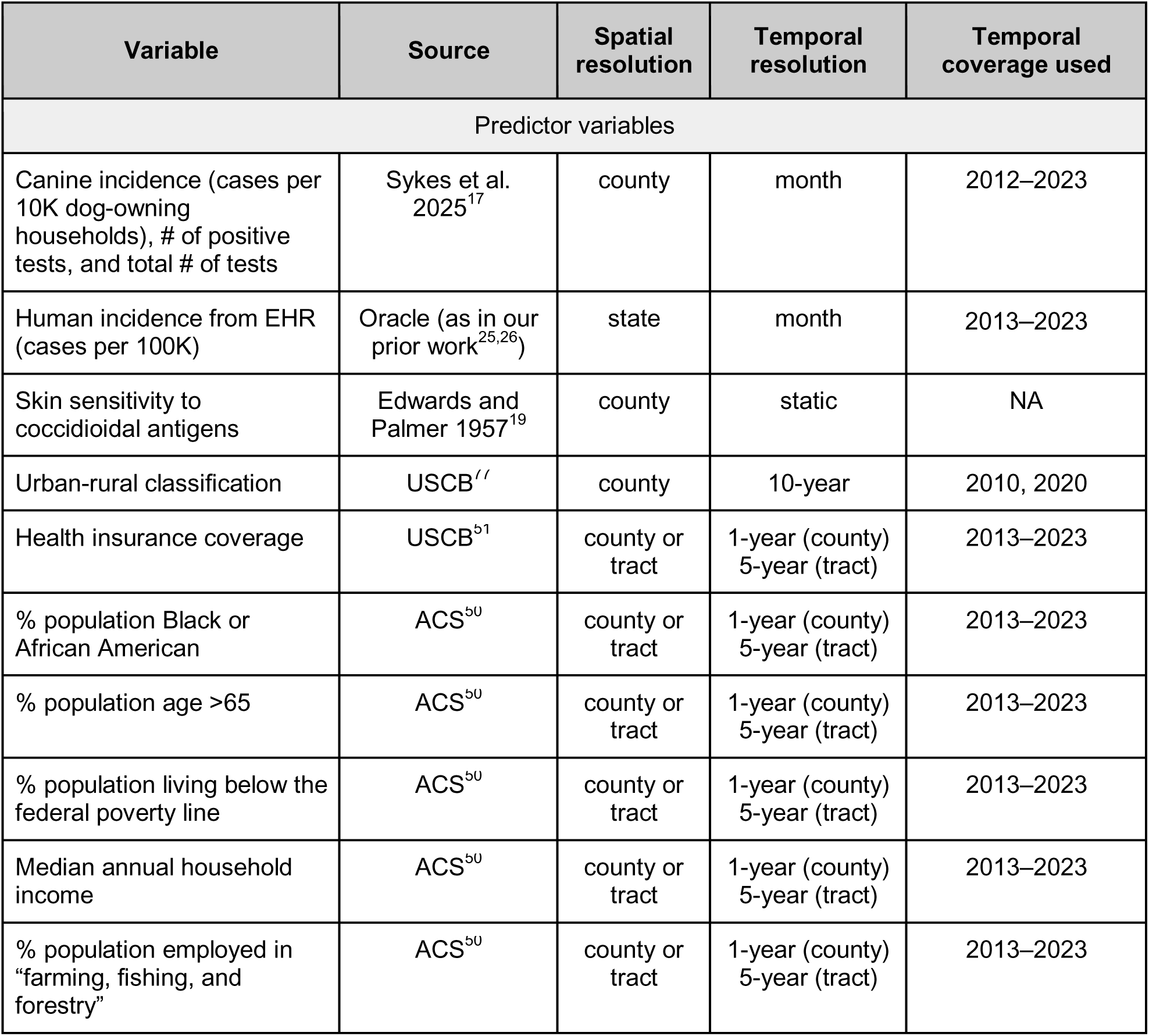
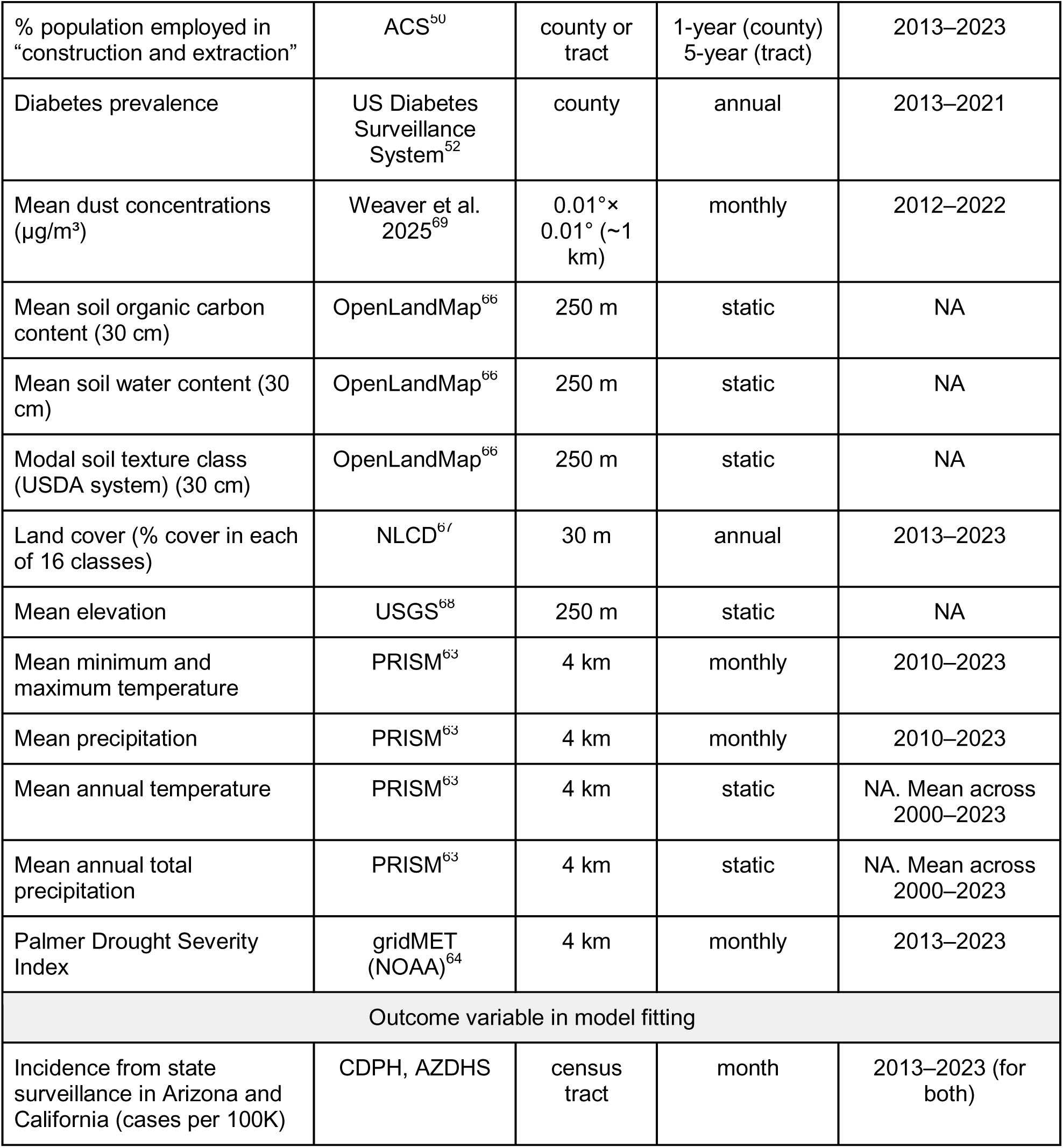
Data used to model US coccidioidomycosis incidence. Summary of predictor and outcome data used in the XGBoost model to estimate monthly, county-level coccidioidomycosis incidence across the US. See eAppendix 1 in Supplement 1 for further details (including case definitions) for each data source. Spatial resolution refers to the native resolution of the datasets—all data were aggregated to the census tract or county level for analysis. Note that we restricted our analysis to 2013–2023, even though several sources have data available before and/or after this period. Data on canine serological testing, dust concentrations, and monthly temperature and precipitation were used starting at earlier time periods as they were included at varying lags in the model.

## Methods

### Overall approach

The outcome of our machine learning model (described below) was monthly, county-level coccidioidomycosis incidence. Specifically, we fit the model to monthly, census tract-level surveillance data from Arizona and California, then applied the model to all other states in the contiguous US to estimate monthly, county-level incidence. Thus, our model predictions represent the coccidioidomycosis incidence that would have been observed if all states had reporting reliability, diagnostic practices, and health care-seeking rates similar to Arizona and California. As predictors, we included a suite of variables serving as proxies for variation in environmental suitability for *Coccidioides*, exposure, disease severity, health care-seeking behavior, and diagnosis and reporting (Table 1, Figure 1).

**Figure 1.**
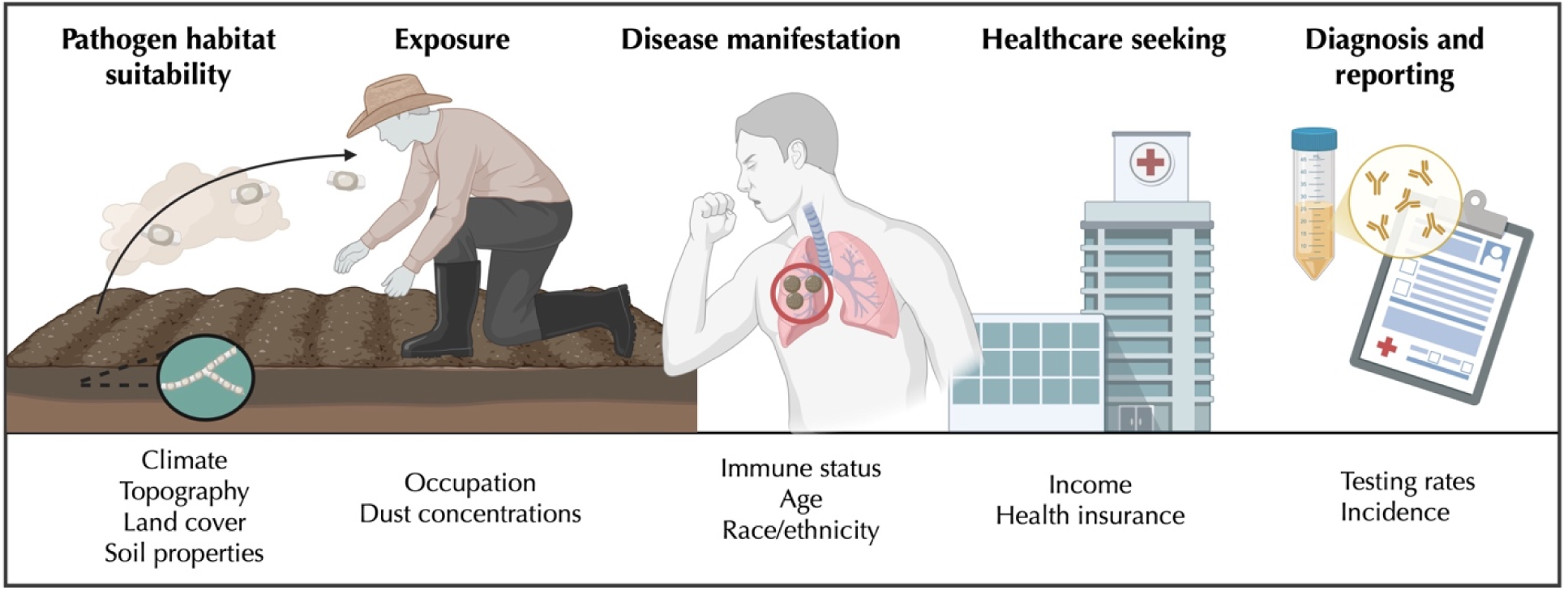
Processes involved in the pathway to a reported coccidioidomycosis case. Factors associated with each process are noted below the diagram (but see Table 1 for the list of specific predictors included in the model including definitions, resolution, and sources).

### Data sources

#### Arizona and California census tract-level surveillance data

The model was fit to monthly, census tract-level data on incident coccidioidomycosis cases from 2013–2023 from state surveillance conducted by the Arizona Department of Health Services (AZDHS) and the California Department of Public Health (CDPH), obtained through data use agreements. Cases were defined on the basis of a positive *Coccidioides* laboratory test and/or clinical criteria, as reported by health care providers and laboratories within these two states (eAppendix 1 in Supplement 1)^42,43^. The month listed for a given case is based on the estimated date of disease onset (estimated symptom onset date if available, otherwise the earliest recorded clinical or laboratory date). Case reports are de-duplicated and only the earliest record is kept. We calculated incidence from these case data using tract-level population estimates from the US Census Bureau (USCB)^44^.

#### Electronic health record data

We obtained deidentified electronic health record data for the period 2013–2023 on patient encounters with an International Classification of Diseases (ICD) 9th or 10th revision code indicative of a coccidioidomycosis diagnosis from Oracle EHR Real World Data (OERWD). OERWD is a longitudinal database containing records of over 110 million unique patients across 176 health systems, fully described in our prior work^25,26^. In this study, we defined cases as patients with a new ICD code indicative of a coccidioidomycosis diagnosis (i.e., ICD-9: 114*; ICD-10: B38*) recorded in the primary diagnostic position, in either a single inpatient visit or two outpatient visits within 30 days. Oracle Health geolocated records at the state level based on patient addresses, and we estimated monthly incidence rates (cases per 100,000 population) using population estimates from the USCB^44^. Because the OERWD database captures only patients receiving care within Oracle Health Systems, we adjusted for differential patient representation relative to the US population by constructing post-stratification weights for the coccidioidomycosis case cohort using American Community Survey (ACS) 5-year population estimates, consistent with our prior work^25^. These monthly, state-level coccidioidomycosis incidence rates were downscaled to the tract or county level based on population size estimates.

#### National skin test data

We obtained data on nationwide prevalence of skin-test reactivity to coccidioidal antigens (“coccidioidin”)^19^. Skin test surveys were conducted on an approximately nationally representative sample of young adults between 1945–1951 (see eAppendix 1 in Supplement 1 for additional details on skin test and study participants). The published, county-level skin test positivity estimates were restricted to white men and women between 17–21 years of age who had not been away from their home county for more than six months (approximately 48,000 individuals). We digitized these skin test positivity estimates from the original study figure and categorized them into the prevalence tiers as reported in that study (0–1%, 1–5%, 6–10%, 10– 30%, 30–50%, and 50–70%).

#### Canine coccidioidomycosis data

We obtained deidentified data on canine coccidioidomycosis cases at the monthly, county level through a data use agreement with the seven major diagnostic laboratories that conduct canine serological testing in the US, as described in our prior work^17^. We defined canine cases as any positive serological test (either IgG or IgM) and calculated canine coccidioidomycosis incidence as the number of cases per 10,000 dog-owning households^17^ using counts of households at the annual, county level from the USCB and estimates of dog ownership in urban and rural communities (35.5% and 54.5%, respectively)^45^ (eAppendix 1 in Supplement 1). We included canine incidence rates, as well as the number of positive serological tests and the total number of serological tests conducted as three separate predictors to capture test positivity and testing effort.

#### National county-level surveillance data

For comparing our model predictions, we obtained monthly, county-level data from 2013–2022 on incident coccidioidomycosis cases for the 28 US states with mandatory reporting from the National Notifiable Disease Surveillance System (NNDSS) operated by the Centers for Disease Control and Prevention (CDC) through a data use agreement. Herein, cases in high-incidence states (Arizona and California) are defined based on confirmatory or presumptive laboratory evidence. Cases in low-incidence states (all other states) are defined based on a) confirmatory laboratory evidence and either epidemiologic linkage (exposure to a *Coccidioides* endemic area) or clinical criteria, or b) presumptive laboratory evidence, epidemiologic linkage, and clinical criteria^33^. See eAppendix 1 in Supplement 1 for additional details regarding clinical and laboratory criteria, and epidemiologic linkage. As with the state surveillance data, we calculated incidence using population estimates from the USCB^44^.

#### Sociodemographic data sources

We included a suite of sociodemographic predictors as proxies for variation in exposure, disease severity, and human health care-seeking behavior, based on prior evidence^46–49^. These included the percentage of the population employed in “construction and extraction” or “farming, fishing, and forestry”; the percentage aged >65 years; the percentage identifying as Black or African American; the percentage living below the federal poverty line; the percentage uninsured (and <65 years); and the median household income. These variables were defined using tract-level estimates from the ACS^50^ and the Small Area Health Insurance Estimates Program of the USCB^51^. We also included the prevalence of diabetes using data from the CDC’s US Diabetes Surveillance System^52^. As the diabetes data were only available at the county level, we applied prevalence values uniformly to all census tracts within a given county when training and testing the model. Further, we obtained population size estimates at the annual, tract level for Arizona and California from the Arizona Office of Economic Opportunity and the California Department of Finance, and at the annual, county level for all states from the USCB^44,53,54^.

#### Ecological data sources

We included ecological predictors in the model based on prior evidence that the distribution and life cycle of *Coccidioides* are influenced by climate conditions^11,55–57^, physical and chemical soil properties^11,28–30,58^, and land cover^30,59–61^. Specifically, we included monthly minimum and maximum temperature and total precipitation, each at a 2-to 26-month lag, following our prior work^62^. We calculated tract- or county-level values of these variables using 4 km resolution data from the Precipitation-elevation Regressions on Independent Slopes Model (PRISM)^63^. We used this same data source to define static variables for mean temperature and total precipitation averaged across 2000–2023. We additionally included monthly estimates of the Palmer Drought Severity Index (PDSI) using the 4 km resolution gridMET dataset^64^.

For soil properties, we included organic carbon content, water content, and soil texture (USDA classification system^65^) at 30 cm depth, using estimates from the OpenLandMap dataset^66^. These soil property data were available at a 250 m spatial resolution and represent time-invariant baseline conditions. For land cover, we included the proportion of cover in each of 16 classes (eAppendix 1 in Supplement 1), using annual, 30 m resolution data from the National Land Cover Database^67^. We also included the mean elevation of each region, using the 250 m resolution digital elevation model from the US Geological Survey^68^. Lastly, we included monthly fine mineral dust concentrations (µg/m³) at 0.01°× 0.01° (approximately 1 km) resolution following our prior work^69,70^.

### Model specification and fitting

We modeled monthly, county-level coccidioidomycosis incidence using extreme gradient boosting (XGBoost), a machine learning algorithm wherein weak decision trees are sequentially built from the prediction errors of the prior tree, resulting in a strong ensemble prediction model^71^. This approach has been demonstrated to achieve high predictive accuracy while minimizing overfitting, allows for nonlinear relationships among predictors and outcome, and is robust to high multicollinearity among predictors^71–73^. We fit the model to monthly, census tract-level surveillance data from Arizona and California using a Tweedie regression objective, as the data exhibited a zero-inflated and right-skewed distribution^74^ (eFigure 1 in Supplement 1). We excluded any tracts with fewer than 100 people as incidence data and socioecological predictors expressed as proportions were highly unstable.

We tuned hyperparameters controlling model complexity and overfitting using a Bayesian optimization approach with five-fold cross-validation implemented with *rBayesianOptimization*^75^ (eAppendix 1 and eTable 1 in Supplement 1). We ran the final model using the optimal hyperparameters, and an 80/20% random split of census tracts into training and testing subsets, respectively. To account for variation that may arise from randomly sampling training and testing subsets, and to get an estimate of model uncertainty, we repeated the final model fitting procedure 100 times, randomly drawing a new 80% training set each iteration. This bootstrapping approach enabled us to obtain distributions of model predictions, performance metrics, and predictor importance values. When reporting uncertainty, values are based on the 2.5^th^ and 97.5^th^ percentile of the bootstrap distribution. To assess the sensitivity of our results to the choice of model predictors, we ran bootstrapped models (b = 10) including either a) all predictors except the veterinary and epidemiological predictors (canine serology and EHR-based incidence), or b) all predictors except coccidioidin reactivity.

We evaluated model predictive accuracy by comparing the predicted and reported cases for Arizona and California and calculating R^2^, RMSE, and Tweedie deviance. We compared our national county-level incidence rate predictions to incidence rates calculated based on the CDC’s NNDSS, which contains monthly, county-level data for all states with mandatory reporting (n = 28)^10^. We evaluated feature importance during each of the bootstrap iterations by calculating Shapley Additive Explanation (SHAP) values, which quantify the marginal contribution of a given feature to model predictions across all possible combinations of the remaining features^76^. We examined partial dependence plots of the relationship between model predictions and a given feature.

### Ethical approval

The analyses involving human subjects’ data received approval from the Committee for Protection of Human Subjects of the California Health and Human Services Agency (protocol no. 17-05-2993) and the Arizona Department of Health Services Human Subjects Review Board (protocol no. 25-0018). Approval by the University of California, Berkeley was provided by reliance on the California State approval.

## Results

### Model performance

We compared our model’s out-of-sample predictions (averaged across 100 bootstrap iterations) to reported incident cases in Arizona and California and found that predictive accuracy was moderate to very high when evaluated at the tract-month ( = 0.291; n = 1,132,537), tract-year ( = 0.681; n = 94,391), county-month ( = 0.996; n = 9,636) and county-year ( = 0.998; n = 803) levels (see eFigure 2 and eTable 2 in Supplement 1 for additional performance metrics and uncertainty). In general, our model tended to underpredict incident cases at the tract-month level relative to reported incident cases by an average of 11.9% (95% bootstrap interval, −15.2% to −7.9%). Model performance was highly similar under alternative model specifications that excluded either the veterinary and epidemiological predictors or coccidioidin reactivity (eTable 3 in Supplement 1). The lack of a substantial reduction in model performance when excluding these predictors may be because their effects were captured by the remaining socioecological predictors. For example, EHR-based incidence and 3-month lagged dust concentrations were moderately correlated (r = 0.63; averaged across months and tracts), as were coccidioidin reactivity and mean annual temperature (r = 0.67) (eFigure 3 in Supplement 1).

We examined spatial biases in our model by summing the mean out-of-sample predictions in Arizona and California across all months from 2013–2023 for a given tract and computing the extent of under- or overprediction compared to reported case totals. We did not observe any clear spatial pattern in tract-level biases (eFigure 4 in Supplement 1). However, underpredicted tracts (here defined as those for which predicted cases were at least 50% lower than reported cases) typically had lower mean annual temperature, higher soil organic carbon content, and higher mean annual precipitation compared to more accurately predicted tracts, while overpredicted tracts (predicted cases were at least 50% higher than reported cases) typically had larger population sizes (eFigure 5 in Supplement 1). In general, our model recapitulated the seasonal pattern of reported cases in Arizona and California in the past decade (eFigure 6 in Supplement 1). In particular, the predicted cases by month-year more closely matched the observed case time series for Arizona ( = 0.97) than California ( = 0.69), where the model tended to underpredict case counts, particularly in peak periods such as in fall 2017 (eFigure 6 in Supplement 1).

### Nationwide incidence predictions

When applied to all counties across the contiguous US, our model estimated the highest annual incidence in the southwest (Figure 2a). In particular, counties predicted as high-incidence (defined as ≥10 annual cases per 100,000 population on average across 2013–2023 per prior work^32^) were concentrated in Arizona (n = 14 out of 15 total counties), California (n = 17/58), Nevada (n = 2/17), New Mexico (n = 8/33), Texas (n = 8/254), and Utah (n = 1/29) (Figure 2b). Counties predicted as moderate-incidence (defined as 2–10 annual cases per 100,000 population on average across 2013–2023^32^) were located in the states noted above—Arizona (n = 1/15), California (n = 11/58), Nevada (n = 7/17), New Mexico (n = 21/33), Texas (n = 42/254), and Utah (n = 7/29)—as well as in Colorado (n = 20/64), Idaho (n = 9/44), Kansas (n = 2/105), Montana (n = 11/56), Nebraska (n = 3/93), Oklahoma (n = 1/77), Oregon (n = 4/36), Washington (n = 7/39), and Wyoming (n = 5/23) (Figure 2b). For all other states, no county had an estimated mean annual incidence above 2 cases per 100,000 population. Relative uncertainty in predicted incidence was also lowest in the southwestern US (eFigure 7 in Supplement 1).

**Figure 2.**
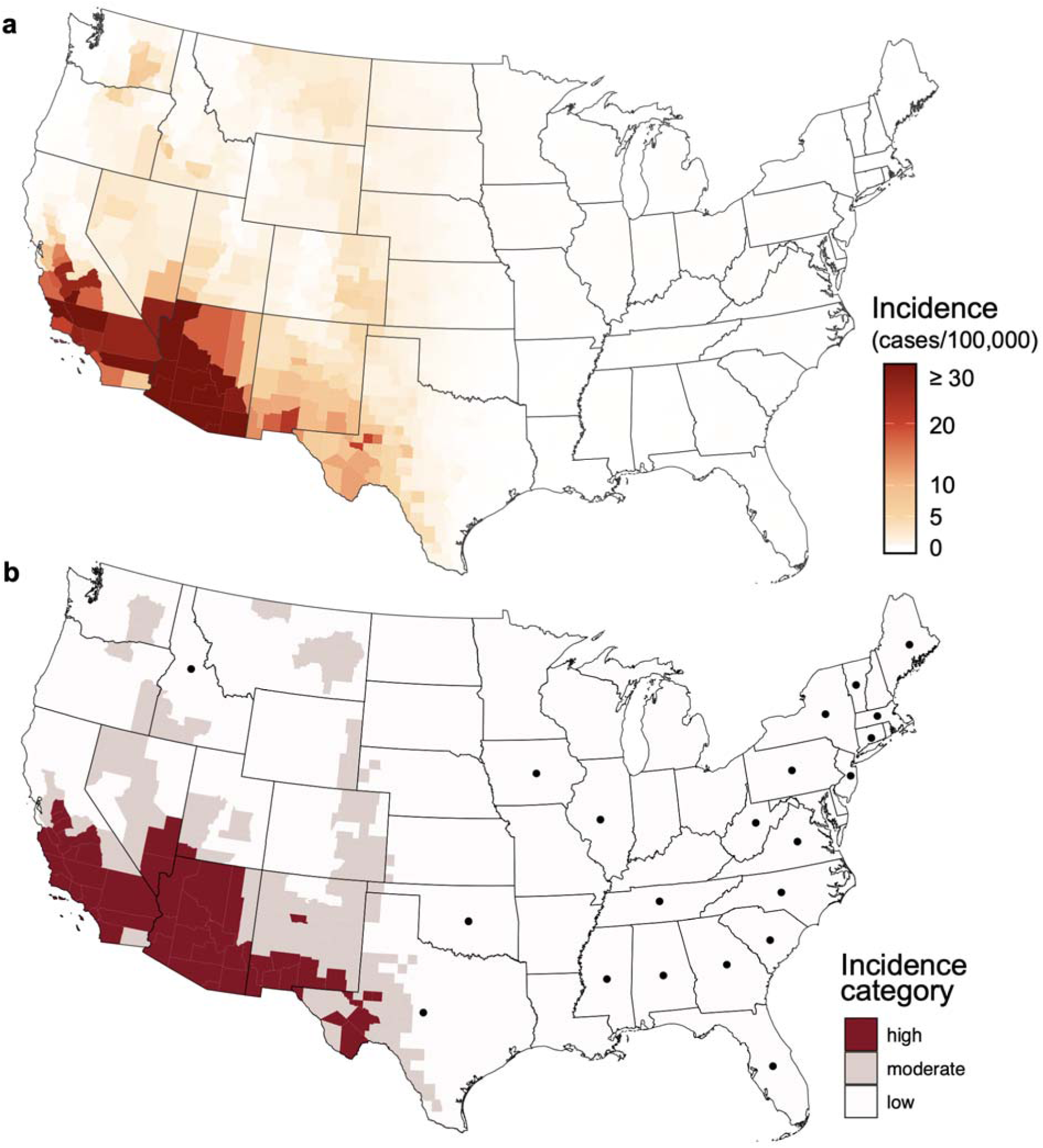
Model-predicted mean annual coccidioidomycosis incidence. Predicted coccidioidomycosis incidence (cases per 100,000 population) across all US counties averaged across all months from 2013–2023. Incidence values in **a)** are shown on a continuous scale, with values capped at 30 to aid visualization. The same underlying data are shown in **b)**, but with incidence values categorized as high (≥10 cases per 100,000 population), moderate (2–10 cases per 100,000 population), or low (<2 cases per 100,000 population). Dots denote states that lack mandatory reporting (n = 22 states as of 2026^18^).

The predicted distribution of coccidioidomycosis incidence was highly similar under the alternative model specifications, remaining constrained to the western US and highest in the southwest. However, under alternative specifications, high-incidence counties were also estimated in Idaho, Colorado, Oregon, and/or Washington, and moderate-incidence counties were also estimated in North and/or South Dakota (eTable 4 and eFigure 8 in Supplement 1).

When aggregating predicted incident cases to the state level, the states with the highest predicted mean annual incidence were Arizona, California, Nevada, and New Mexico (eFigure 9 in Supplement 1). In particular, these states all had annual incidence ≥10 cases per 100,000 population for a period of three or more consecutive years—the classification for “high-incidence jurisdictions” used by the CDC^33^. However, this state-level definition likely masks regional patterns of high incidence, as western Texas (as defined by the 36 southwestern-most counties^34^) would also be classified as high incidence based on our model predictions, while Texas as a whole is not (eFigure 10 in Supplement 1). The states with the highest total predicted incident cases were Arizona, California, Nevada, New Mexico, and Texas (eTable 5 in Supplement 1).

For states outside of Arizona and California that have mandatory reporting, our model predictions were moderately correlated with monthly, county-level incident coccidioidomycosis cases reported to the CDC’s NNDSS ( = 0.60; n = 174,360) (Figure 3, eFigure 11 in Supplement 1). Our model predicted higher county-level incidence in these states by an average of 28% (95% bootstrap interval, 0.1%–69.8%), equating to a total of 608 additional cases per year relative to NNDSS reporting (95% bootstrap interval, 340–964 cases) (see eTable 5 in Supplement 1 for prediction discrepancies by state). The extent to which model predictions differed from NNDSS reporting varied widely by state, ranging from 350% higher than reported incidence in Nevada to 95% lower in Louisiana (eTable 5 in Supplement 1).

**Figure 3.**
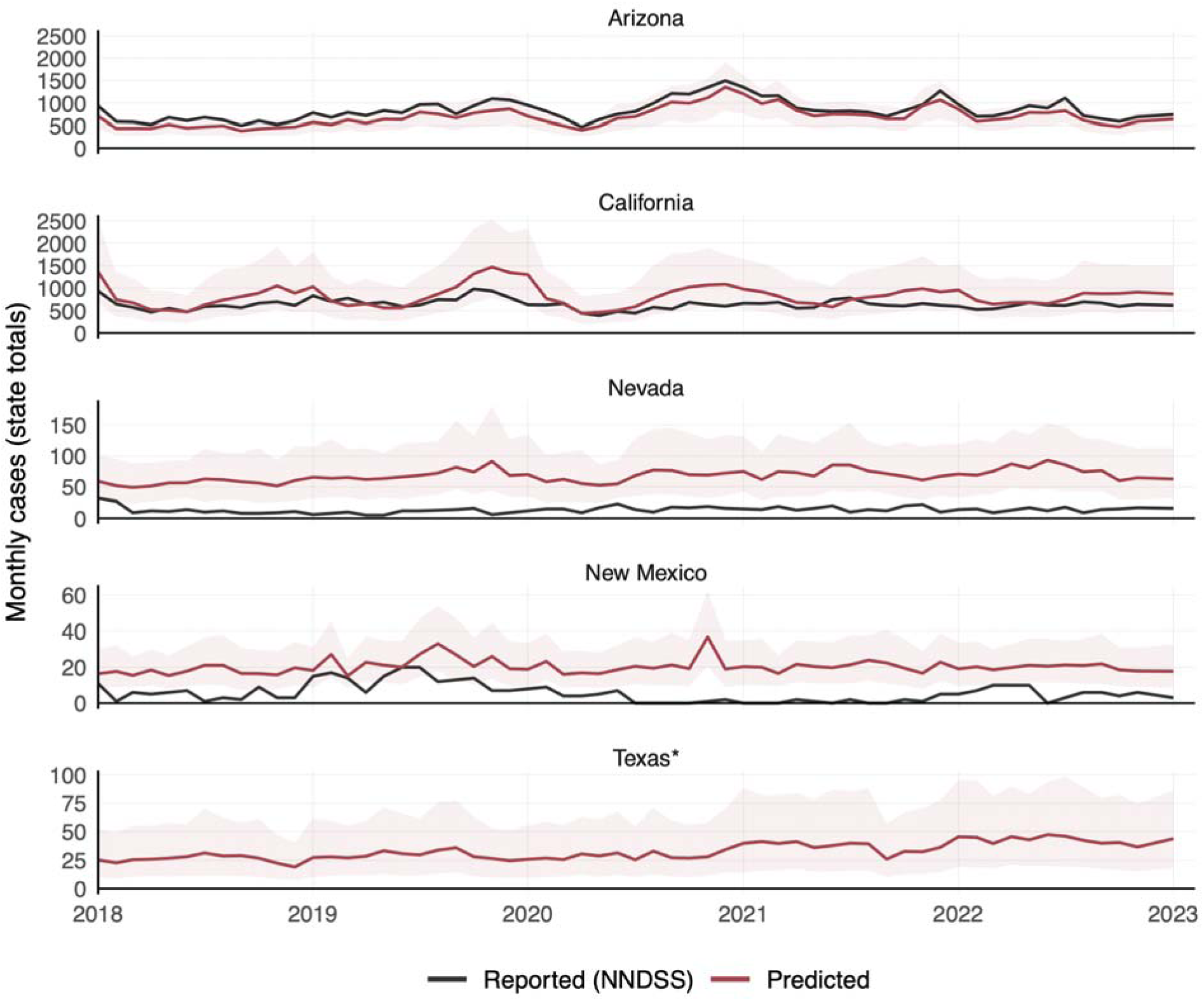
Time series of monthly coccidioidomycosis cases. Monthly coccidioidomycosis cases from January 2018 to December 2022 for select states (the top five in total predicted case counts) based on model predictions (red) and cases reported to the CDC’s NNDSS (black). The (*) denotes a state without mandatory reporting. The red line and light red shading denote the mean and 95% bootstrap interval, respectively, of the model’s out-of-sample predictions across 100 bootstrap iterations. Note that the y-axis limits differ by state.

States where model predictions were lower than reported incidence were typically outside of the suspected region of endemicity for *Coccidioides* (eFigure 12 in Supplement 1); thus, the discrepancy may be due in part to travel-associated exposures. That is, the NNDSS reports cases based on patient residence, which may or may not be where the patient was exposed to *Coccidioides*. Across states with the highest case counts, our model predictions showed similar seasonal timing to that of reported cases, with predicted and reported cases generally peaking in similar months (Figure 3).

We identified 10 counties outside of Arizona and California that would be classified as high incidence based on our model, but low incidence based on NNDSS case reports (compared to 3 counties in which the opposite occurred) (eFigure 13 in Supplement 1). These counties occurred in Nevada and New Mexico and tended to have higher numbers of positive and total canine serological tests, higher EHR-based incidence, and higher scrub-shrub land cover (eFigure 14 in Supplement 1).

### Feature importance

The features contributing the most to model predictions (i.e., those with the highest SHAP values) were canine coccidioidomycosis incidence, the number of positive canine serological tests, mean annual precipitation and temperature, coccidioidin reactivity, elevation, and EHR-based incidence (Figure 4a). Predicted cases increased with increasing canine incidence, positive canine serological tests, coccidioidin reactivity, EHR-based incidence, and mean annual temperature; decreased with increasing mean annual precipitation; and peaked at intermediate elevation (∼500 m) (Figure 4b, eFigure 15 in Supplement 1).

**Figure 4.**
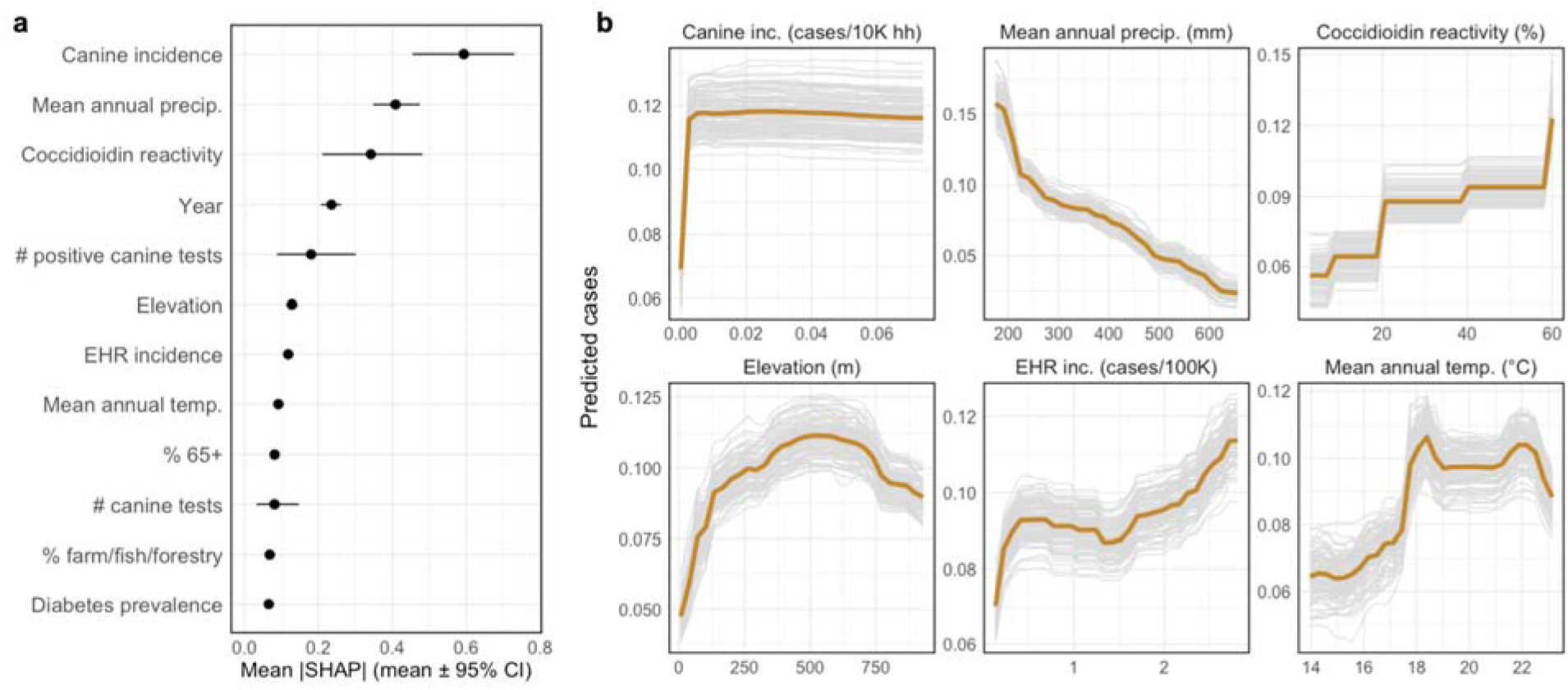
Feature importance. **a)** Feature contribution to coccidioidomycosis case predictions. Points are the mean marginal contribution of a given feature to model predictions across all possible feature combinations (|SHAP| value). Error bars denote the 95% bootstrap interval computed across 100 bootstrap iterations. Only the top 12 predictors are shown here. **b)** Partial dependence plots showing the response of predicted cases to a given feature averaged across all possible feature combinations. The orange line denotes the mean response across model iterations, while the gray lines denote the response from each of 100 bootstrap iterations. Only select features among the most important are shown here: canine incidence, mean annual precipitation, coccidioidin reactivity, elevation, EHR-based incidence, and mean annual temperature.

In the model specifications that excluded either coccidioidin reactivity or the EHR and canine serology predictors, the most important features were mean annual precipitation and temperature, elevation, and the percent of the population employed in “farming, fishing, and forestry” (eFigure 16 in Supplement 1).

## Discussion

Despite rising public health concern over coccidioidomycosis, its distribution and incidence remain poorly characterized given the absence of robust national surveillance. In this study, we integrated epidemiological, veterinary, and socioecological data sources in a machine learning analysis to address gaps in mandatory reporting and limitations in case ascertainment outside the most highly endemic states. We identified regions with elevated incidence in non-reporting states and quantified the extent of case under-ascertainment relative to Arizona and California in states with mandatory reporting. By advancing understanding of which regions may be endemic for coccidioidomycosis, our results may help improve public and physician awareness, supporting more effective disease management and control.

Our results suggest a broader distribution of coccidioidomycosis in the US compared to most prior ecological models of *Coccidioides* spp.^28–31^. In particular, we estimated that high-incidence counties (defined as ≥10 mean annual cases per 100,000 population across 2013–2023, following prior work^32^) are concentrated in the southwest—namely Arizona, California, Nevada, New Mexico, Texas, and Utah (Figure 2). However, moderate-incidence counties (2–10 mean annual cases per 100,000 population across 2013–2023^32^) occur within several additional states, namely Colorado, Idaho, Kansas, Montana, Nebraska, Oklahoma, Oregon, Washington, and Wyoming (Figure 2b). Notably, our predicted distribution of high- or moderate-incidence counties aligns closely with the potential range of *Coccidioides* estimated by the CDC, with the exception of western North and South Dakota, which the CDC includes in that range^35^ but where we predict low incidence.

Our model identified 60 high- or moderate-incidence counties within states without mandatory reporting of coccidioidomycosis, including nine in Idaho (moderate), one in Oklahoma (moderate), and 50 in Texas (8 high, 42 moderate), underscoring gaps in data and understanding that stem from the absence of nationwide mandatory reporting (Figure 2).

Further, for states outside of Arizona and California that have mandatory reporting, we identified 10 counties that would be classified as high-incidence based on our model, but low-incidence based on cases currently being reported to CDC’s NNDSS (eFigure 13 in Supplement 1). For these states, we predicted annual, county-level incidence rates that were 28% higher, on average, than those reported to NNDSS, equating to a total of 608 additional cases per year. These discrepancies are likely in part because our predictions represent a counterfactual scenario in which all regions had similar rates of health care seeking for uncomplicated disease, accurate diagnosis, and case reporting for coccidioidomycosis as those of Arizona and California—presumed to have the highest rates of case ascertainment in the US.^6^ Thus these results suggest that case under-ascertainment in states outside the highly endemic region— even where reporting is mandatory—may hinder identification of high-incidence counties.

At the state level, our model identified Arizona, California, Nevada, and New Mexico as meeting the CDC criteria for “high-incidence jurisdictions”, defined as having annual incidence ≥10 cases per 100,000 population for a period of three or more consecutive years^33^. Notably, this expands upon the states considered high-incidence jurisdictions, which, as of July 2022, were restricted to Arizona and California^33^. While Texas as a whole did not meet the “high-incidence jurisdiction” criteria given low incidence rates in the eastern half of the state, our predictions identified western Texas—a region with approximately 1.5 million people—as exceeding the threshold (eFigure 10 in Supplement 1). This finding aligns with prior work identifying western Texas as highly endemic for coccidioidomycosis^36–38^.

Of the features included in our model, we found that the epidemiological and veterinary predictors, as well as mean annual precipitation, had the largest influence on model predictions (Figure 4). As expected, monthly incidence predictions in Arizona and California increased with higher canine incidence and positive canine serological tests, as well as higher human incidence as ascertained from electronic health records. These findings align with prior work reporting strong correlations between canine and human incidence in Arizona and California^17^, and highlight the benefit of multisource epidemiological data for characterizing disease burdens in the absence of robust national surveillance. Predicted incidence also increased with higher levels of coccidioidin reactivity—a predictor based on large-scale skin test surveys conducted in the 1940s and 1950s^19^—suggesting that this initial assessment still provides information regarding the current spatial distribution of the disease. Predicted incidence also decreased steadily with increasing mean annual precipitation, with very few predicted cases occurring above 600 mm, aligning with prior work finding higher incidence of coccidioidomycosis under drier conditions in the southwestern US^15^, and associations between *Coccidioides* and arid soils^11,29^.

Our modeling approach has several limitations that warrant acknowledgement. First, our incidence estimates do not distinguish between local versus travel-acquired infections, as these are not distinguished in the EHR or canine serological data used in our model (and are also not distinguished in the NNDSS wherein cases are reported based on the patients’ residence).

However, our model predicted low incidence across eastern states presumed to be non-endemic for *Coccidioides* despite high canine test positivity in locations such as Florida and Maine, suggesting we may have minimized the influence of travel-acquired infections by incorporating predictors associated with local pathogen presence (e.g., coccidioidin reactivity, baseline and monthly climate conditions, soil properties). Second, reported coccidioidomycosis cases are zero-inflated (i.e., approximately 91% of tract-months had zero reported cases), which makes accurate model prediction challenging^39^ and can lead to wide bootstrap intervals, even though we used a compound Poisson-gamma distribution (Tweedie distribution) designed for zero-inflated, right-skewed data. Third, while our predictor data sources have national coverage, there may be biases in their comprehensiveness. For example, the coccidioidin skin test surveys were conducted only on white men and women between 17–21 years of age; and the EHR data used here include only data on health systems with a data use agreement with Oracle (although we did adjust for differential subgroup representation here using post-stratification weighting as in prior work^25,26^).

Additionally, our model is not designed to capture or replicate outbreaks such as those driven by natural disasters (e.g., dust storms, earthquakes) or specific occupational exposures (e.g., construction, wildland firefighting). That is, while our model includes monthly climate and dust predictors, as well as annual occupation predictors, these better capture general suitability for pathogen presence and exposure in a given region and time rather than specific exposure events. Further, localized outbreaks may not be well reflected in the state-level EHR data, and occupation-related outbreaks would not be reflected in higher canine seropositivity. Lastly, despite fitting our model to surveillance data from Arizona and California—wherein rates of case ascertainment are expected to be the highest^6,10^—under-ascertainment, underreporting, and misdiagnosis are still inherent and geographically variable in our outcome data. Thus, our model does not capture the total coccidioidomycosis incidence for a given region, but rather the expected incidence assuming similar rates of case capture as Arizona and California.

Our model provides an up-to-date and comprehensive estimate of the monthly distribution of coccidioidomycosis incidence across the US. Improved estimates of disease incidence— particularly those that minimize gaps due to case under-ascertainment and that are available across all US states regardless of reporting requirements—may help improve public and physician awareness, supporting more timely and accurate diagnoses. In particular, patients with knowledge of coccidioidomycosis have been found to receive earlier diagnoses and be more likely to request testing for coccidioidomycosis from their health care provider^40^. Similarly, an intervention aimed at increasing awareness among emergency medicine providers was found to increase testing for coccidioidomycosis in patients with suspected community-acquired pneumonia (of which *Coccidioides* is a common cause in endemic regions)^41^. More broadly, an improved understanding of the nationwide coccidioidomycosis distribution is critical for detecting and preparing for emergence in new regions and understanding how disease burdens may shift in the coming decades under environmental and societal changes.

## Author contributions

LIC, SKC, JES, and JVR conceived of the research study. LIC, SKC, XL, AEH, BLMB, AKH, JRH, and JVR designed the analysis. LIC, SKC, JES, BLMB, GSC, TW, IR, and AKH provided and/or compiled the data. LIC conducted the formal analysis. All authors received and edited the final draft and approved of submission.

## Data Availability

All data produced in the present study are available upon reasonable request to the authors.

