## Supplementary Information for "National incidence of coccidioidomycosis in the United States estimated through machine learning integration of epidemiological, veterinary, and socioecological data"

**eAppendix 1. Supplementary Methods**

**eFigure 1.** Histogram of tract-level incidence

**eFigure 2.** Model performance for Arizona and California census tracts

**eFigure 3.** Predictor correlation matrix

**eFigure 4.** Spatial bias by census tract

**eFigure 5.** Features of over- or underpredicted census tracts

**eFigure 6.** Predicted and reported monthly case counts

**eFigure 7.** Relative uncertainty in predicted annual incidence

**eFigure 8.** Nationwide incidence predictions under alternative model specifications

**eFigure 9**. Predicted state-level mean annual incidence

**eFigure 10.** Mean annual incidence in western Texas

**eFigure 11.** Predicted versus NNDSS reported cases

**eFigure 12.** Extent of over- or underprediction by state

**eFigure 13.** Discrepancies in counties considered high-incidence

**eFigure 14.** Features of counties predicted as high-incidence

**eFigure 15.** Partial dependence plots for additional model predictors

**eFigure 16.** Feature importance in alternative model specifications

**eTable 1.** Model hyperparameters tuned using Bayesian optimization

**eTable 2.** Additional model performance metrics

**eTable 3**. Performance metrics under various model specifications

**eTable 4.** Incidence predictions under various model specifications

**eTable 5.** State-level incidence and cases from model predictions and NNDSS

**eReferences**

**eAppendix 1. Supplementary Methods**

*Arizona and California census tract-level surveillance data*

Monthly, census tract-level surveillance data on incident coccidioidomycosis cases were obtained from the Arizona Department of Health Services (AZDHS) and the California Department of Public Health (CDPH) for the period 2013–2023. These data did not include cases among tribal residents, institutionalized persons, or those without recorded addresses. Cases are only recorded once; thus, patients with more than one reported positive *Coccidioides* test are only recorded upon their initial positive result. For the AZDHS, the laboratory-only case definition (i.e., no clinical criteria required) has been in use since 2008^1^. The only exception is the coccidioidal skin test (conversion from negative to positive), which requires the additional presence of symptoms to count a case as confirmed. However, this was changed in 2022 such that even the coccidioidal skin test alone could be used to define a case^1^. For the CDPH, case classification through the end of 2018 required both a *Coccidioides* positive laboratory test and clinical criteria to be met^2^. Since January 2019, a laboratory-only case definition has been in use^2^.

*Canine coccidioidomycosis data*

Data on canine coccidioidomycosis cases were obtained through a data use agreement with the seven major diagnostic laboratories that conduct canine serological testing in the US (as originally reported in Sykes et al. 2025^3^). To our knowledge, these laboratories account for all canine coccidioidomycosis testing in the US. The data set spans 2012–2023 and includes line-level information on dog age, sex, breed, month and year of testing, county of owners’ residence and/or clinic, test type (agar gel immunodiffusion, complement fixation, or enzyme immunoassay), and result (positive or negative for IgG or IgM). We defined cases as any positive serological test (either IgG or IgM), as in our prior work^3^. Testing and results are only included once for a given dog (i.e., no serial testing). We assigned cases to the owner’s county of residence when available (85.8% of cases) and otherwise assigned cases to the reporting clinics’ county. We calculated canine coccidioidomycosis incidence as the number of cases per 10,000 dog-owning households using counts of households at the annual, county level from the US Census Bureau (USCB)^4^, and adjusting for variation in rates of dog ownership in rural vs urban communities (estimated at 54.5% and 35.5%, respectively)^5^. Specifically, we calculated incidence for a given county and month as:

$$Canine incidence = \frac{\# positive tests}{\# households \times(\% urban pop. \times0.355 + \% rural pop. \times0.545)}$$

We calculated the percent of the county population in urban vs rural communities using a linear extrapolation from the USCB urban-rural classification estimates for 2010 and 2020^6^. To obtain tract-level estimates of these canine serological predictors when fitting the model, we downscaled estimates based on population size distributions (i.e., based on the percent of the county population within a given census tract).

*National county-level surveillance data*

For comparing our model predictions, we obtained monthly, county-level data on incident coccidioidomycosis cases for the 28 US states with mandatory reporting from the CDC’s National Notifiable Disease Surveillance System (NNDSS) through a data use agreement. Herein, cases in high-incidence regions (Arizona and California) are defined on the basis of confirmatory or presumptive laboratory evidence^7^. Cases in all low-incidence regions (all other states) are defined on the basis of a) confirmatory laboratory evidence and either epidemiologic linkage (exposure to a *Coccidioides* endemic area) or clinical criteria, or b) presumptive laboratory evidence, epidemiologic linkage, and clinical criteria^7^.

Confirmatory laboratory evidence consists of: a) positive culture of *Coccidioides* spp., b) histopathological or cytopathological identification of characteristic *Coccidioides* spp. in tissue or body fluid, c) detection of *Coccidioides-*specific nucleic acid or protein sequences in a clinical specimen through a validated molecular assay, or d) detection of coccidioidal antibodies in cerebrospinal fluid, serum, or other body fluids using immunodiffusion, complement fixation (with a titer of >1:2), tube precipitin, or ELISA (detecting both IgM and IgG)^7^. Presumptive laboratory evidence consists of: a) detection of *Coccidioides* spp. antigen in serum, urine, CSF or other body fluids, or b) detection of coccidioidal antibodies in serum or other body fluids using complement fixation (with a titer of 1:2), lateral flow assay, latex agglutination, or ELISA (detecting either IgM or IgG)^7^.

Clinical criteria (in the absence of a more likely diagnosis of an alternative fungal infection) consist of: a) acute onset or worsening of at least two of the following symptoms: cough, fever, chills, night sweats, shortness of breath, chest or flank pain, headache, unintentional weight loss, myalgia, arthralgia or bone pain, or fatigue; or b) at least one of the following findings: abnormal lung findings on chest imaging, single or multiple skin lesions, bone or joint abnormality, meningitis, encephalitis, or focal brain lesion, abscess, granuloma, or lesion in other body system, or erythema nodosum or erythema multiforme rash^7^.

Epidemiologic linkage refers to exposure to a *Coccidioides* spp. endemic area including through residence, travel, or work in the two months prior to acute symptom onset or positive coccidioidal laboratory test if the date of acute symptom onset is unknown. The endemic region for *Coccidioides* is considered by the CDC to include the entirety or portions of the following 12 states: Arizona, California, Colorado, Idaho, Nevada, New Mexico, Oklahoma, Oregon, Texas, Utah, Washington, and Wyoming. If the exposure history is not available, the case is assumed to not meet the epidemiologic linkage criteria.

*National skin test data*

We obtained data on the prevalence of skin-test reactivity to coccidioidal antigens (“coccidioidin”)^8^. These data were gathered on a putatively nationally representative sample of approximately 110,000 young adults between 1945–1951. Namely, skin test surveys were conducted on: 1) young men (n ~ 88,000) as they entered the US Naval Training Center at San Diego, California, which drew recruits from across the US except for the North Atlantic and Great Lakes region; 2) students (n ~ 4,000) at universities in the Great Lakes region and colleges in New England; and 3) female student nurses (n ~ 17,000) at training schools located in metropolitan areas distributed across the US. For each set of surveys, tests were administered via intradermal injection of 0.1 mL of a 1:100 dilution of coccidioidin. Reactions were read at 48–96 hours by measuring the transverse diameter of induration (firm, thickened area of the skin). Each participant was asked to list every city or county in which they lived for more than six months, from birth to the time of testing. The digitized survey results that we used were then restricted to white men and women between 17–21 years of age who had not been away from their home county for more than six months (n ~ 48,000).

The original data on coccidioidin reactivity were mapped at the county level in the following bins: 0–1%, 1–5%, 6–10%, 10–30%, 30–50%, and 50–70%. We digitized this map and defined a numerical coccidioidin reactivity predictor as the midpoint of each bin (i.e., 0.5, 3, 8, 20, 40, and 60). This approach avoids issues with categorical levels that are absent in the training and testing dataset. That is, while many counties across the US were classified as having 0–1% coccidioidin reactivity, this categorical level does not occur for any counties within Arizona and California, thus it would never be learned during model training. Representing reactivity as a continuous numerical variable instead enables the model to generalize to counties with levels that were unobserved during model training.

*Sociodemographic predictors*

We obtained data on the following sociodemographic variables from the USCB American Community Survey (ACS) program: the percent of the population self-identifying as Black or African American; the percent of the population over 65 years of age; the percent of the population living below the federal poverty line; the percent of the population employed in “construction and extraction”; the percent of the population employed in “farming, fishing, and forestry”; and the median household income. Additionally, we obtained data on the percent of the population under the age of 65 without health insurance coverage from the USCB Small Area Health Insurance Estimates Program (SAHIE)^9^. These data are available annually across the study period (2013–2023), at either the annual county level or at the 5-year tract level. We accessed these data using the R package *tidyCensus*^10^. Further, we obtained data on the total number of adults diagnosed with diabetes (as opposed to newly diagnosed; primarily Type 2, but not specifically disaggregated herein) from the CDC’s US Diabetes Surveillance System^11^. These data are available annually at the county level for the period 2004–2021. In our model, we included diabetes prevalence as a percent of the county population. When training the model at the census tract level, we applied the county-level prevalence values uniformly to all census tracts therein.

*Population size estimates*

County-level data on population size were obtained from the USCB and were available at an annual resolution across the study period (2013–2023). To estimate annual tract-level population sizes for Arizona and California we first obtained data on annual county-level population sizes from the Arizona Office of Economic Opportunity^12^ and the California Department of Finance^13^. We then downscaled these estimates based on the proportion of a county’s population size in each census tract as estimated in the 2010 USCB decennial census.

*Dust concentrations*

We utilized data on fine mineral dust concentrations (µg/m³) that our group previously developed^14^. These estimates were derived using a global model of $\mathrm{PM}_{2.5}$ mass and chemical composition developed by van Donkelaar et al. 2019^15^, which uses a chemical transport model (GEOS-Chem), satellite observations of aerosol optical depth, and ground-based observations. Dust estimates had a monthly resolution and 0.01°× 0.01° (approximately 1 km), which we spatially aggregated to the census tract or county level, and used for the period of 2012–2022.

*Soil properties*

We obtained data on the following soil properties from the OpenLandMap dataset^16^: organic carbon content, water content, and soil texture (USDA classification system^17^), each at a 30 cm depth. These global soil property layers are derived from spatiotemporal machine learning models trained on compiled soil profile data and are produced at a 250 m spatial resolution. We accessed these data layers using Google Earth Engine, and aggregated values at the census tract or county level, using means for organic carbon and water content, and the modal soil texture class in each region. These soil property datasets represent long-term baseline conditions and are thus included as static predictors in our model.

*Land cover*

We obtained data on annual land cover classifications from the National Land Cover Database produced by the Multi-Resolution Land Characteristics Consortium^18^. The NLCD provides 30 m resolution pixels from the remotely sensed Landsat data, categorized into one of 16 classes: open water, perennial ice/snow, developed (open space; and low, medium, or high intensity), barren land, forest (deciduous, evergreen, or mixed), shrub/scrub, grassland/herbaceous, pasture/hay, cultivated crops, woody wetland, and emergent herbaceous wetland. These data are updated annually and are available across the study period (2013–2023). As predictors in the model, we used the percent cover of a given census tract or county by each land cover class, calculated using QGIS. Additionally, we included the mean elevation for each tract or county as a static variable, using the US Geological Survey Global Multi-resolution Terrain Elevation Data, available at a 250 m resolution^19^.

*Climate*

We obtained data on mean monthly minimum and maximum temperature and total precipitation across the study period (2013–2023) from the Precipitation-elevation Regressions on Independent Slopes Model (PRISM)^20^. PRISM provides gridded climate estimates for the contiguous US at a 4 km resolution, generated by statistically downscaling observations from a network of monitoring stations^21^. We aggregated gridded estimates at the census tract or county level using QGIS. As predictors in our model, we included each climate variable lagged from 2–26 months, following our prior work identifying lagged effects of climate variables on coccidioidomycosis incidence^22^. To capture long-term climatic suitability, we used the PRISM data to calculate mean annual temperature and mean annual total precipitation averaged across 2000–2023 for a given tract or county, and included these as static predictors. Additionally, we obtained monthly data on the Palmer Drought Severity Index (PDSI)—a standardized index of relative dryness based on temperature, precipitation, and soil-water holding capacity—at a 4 km resolution through the Gridded Surface Meteorological (gridMET) dataset from the National Oceanic and Atmospheric Administration^23^.

*Model specification and fitting*

We modeled monthly coccidioidomycosis incidence using a tree-based machine learning model, extreme gradient boosting (XGBoost). Herein, weak decision trees are sequentially built on the residuals of the prior tree and are combined in an ensemble prediction^24^. This approach has been found to achieve high predictive accuracy across a variety of applications^24–26^, while minimizing overfitting and enabling complex, nonlinear relationships between model predictors and outcome. Herein, we trained and tested the model at the monthly, census tract level for Arizona and California, using the surveillance data and predictors described above. We then applied the final model to estimate incidence across the contiguous US at the monthly, county level. We used XGBoost’s built-in approach for handling missingness in predictor data, wherein branch directions for missing values are learned during training, as this has been found to yield comparable accuracy to pre-processing imputation^27^. As our model objective function, we used a Tweedie regression—a family of distributions for modeling continuous data with a point mass at zero and right-skewed non-zero values^28^—with power parameter (p) of 1.3, as this maximized the log-likelihood of a Tweedie GLM fit to our data.

We tuned model hyperparameters controlling model complexity using Bayesian optimization implemented with *rBayesianOptimization*^29^. Herein, candidate sets of hyperparameter values are iteratively evaluated (here, based on negative log-likelihood) and the next set is selected based on prior iterations. For each hyperparameter, we provided a range of possible values aimed at minimizing overfitting (eTable 1) and set the algorithm to evaluate eight initial random sets (“init_points” = 8), followed by 10 sequential optimization sets (“n_iter” = 10). We used this approach to tune the following model hyperparameters: “eta” (the learning rate), “max_depth” (the maximum tree depth), “gamma” (the minimum loss reduction required to make a tree split), “min_child_weight” (the minimum number of samples required to make a tree split), “subsample” (the proportion of the training data randomly sampled when building each new tree), and “colsample_bytree” (the proportion of available features randomly sampled when building each new tree). For each hyperparameter evaluation, we trained an XGBoost model with “early_stopping_rounds” set to 50, thereby stopping boosting if the evaluation metric did not improve for 50 rounds. We used five-fold cross validation during hyperparameter tuning, with random sampling into folds done based on unique census tract IDs, such that all observations from a given census tract were kept together and fully withheld in at least one fold to improve generalizability (i.e., to avoid leakage of tract-month combinations that share static covariates such as elevation).

We then used the optimal hyperparameters (eTable 1) to fit a final model. Herein, data were randomly split into 80/20% training/testing subsets, again with random sampling done on unique census tract IDs. While all other features had an equal probability of being included in the feature subset for a given tree, we altered the sampling probability applied to the coccidioidin reactivity predictor using “feature_weights” = 0.2, given lower confidence in this dataset (i.e., skin test surveys were conducted several decades ago and the resulting data were of low resolution).

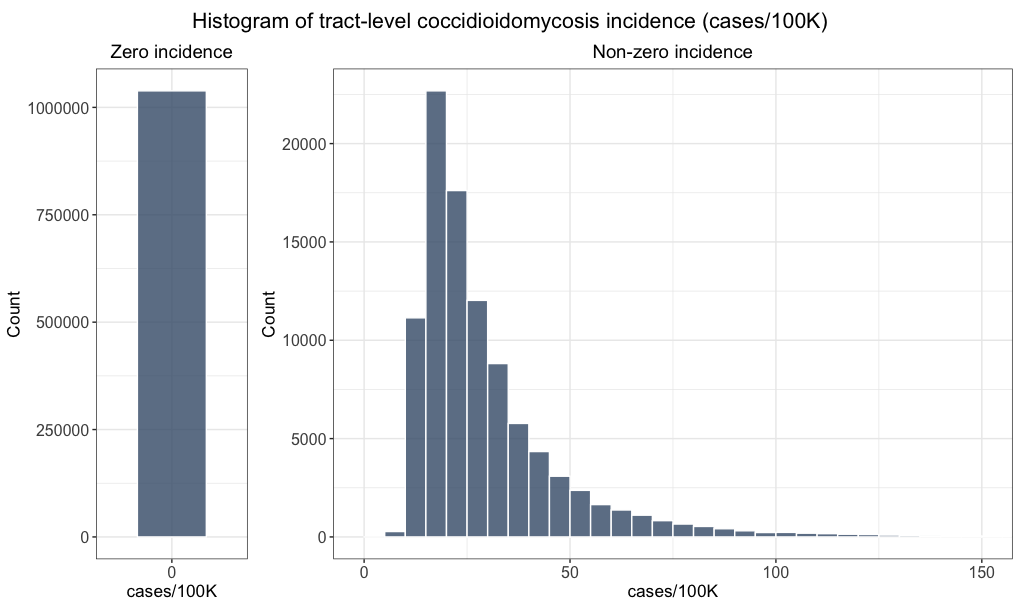

**eFigure 1.** **Histogram of tract-level incidence.** Distribution of coccidioidomycosis incidence (cases per 100,000 population) across Arizona and California census tracts and month-years from 2013–2023 (n = 1,135,309 tract-months). Zero and non-zero incidence are shown on separate plots with different y-axes to aid visualization of variation at the low and high end.

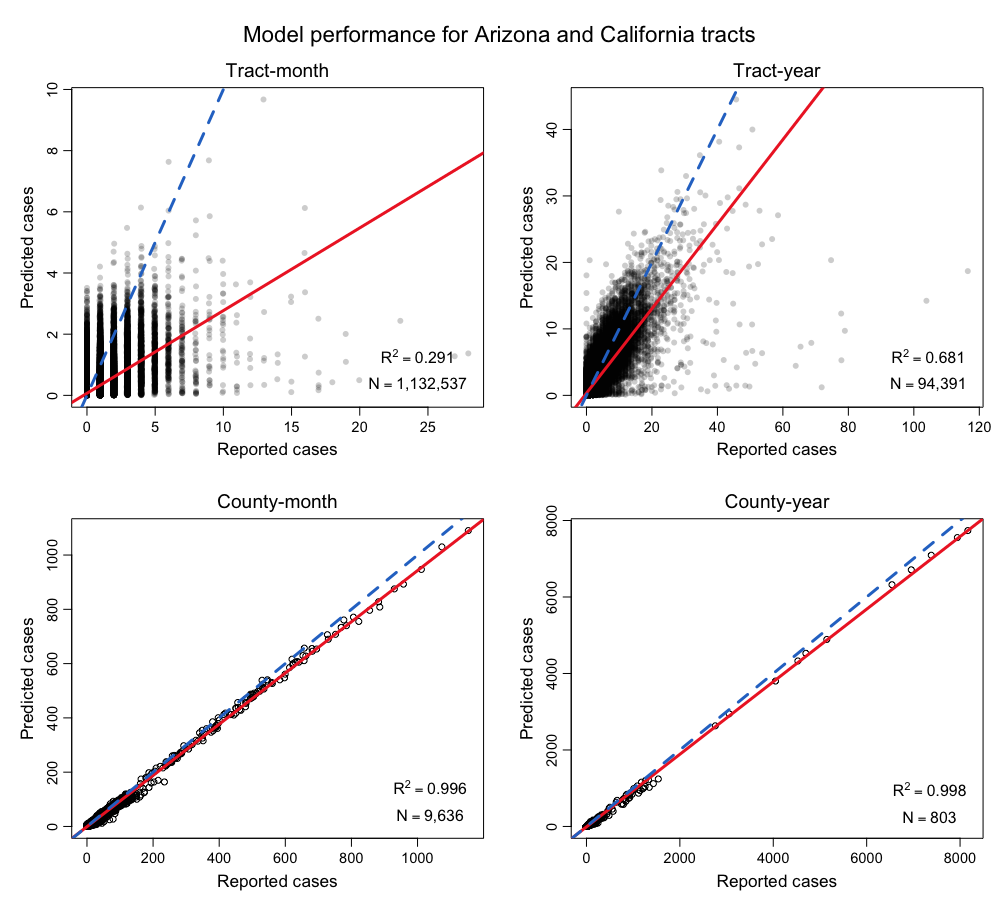

**eFigure 2**. **Model performance for Arizona and California census tracts**. Scatterplot of predicted versus reported coccidioidomycosis cases in Arizona and California at the census tract (top) and county (bottom) level and at the monthly (left) or annual (right) level. Each point denotes the cases for a single tract or county and month or year between 2013–2023. Predicted cases here are the mean across 100 bootstrap iterations. Reported cases are those captured by state surveillance. The solid red line in each plot denotes the regression of predicted against reported cases, while the blue dotted line denotes a 1:1 relationship. The inset text on each plot denotes the $R^{2}$ between predicted and reported cases and the number of observations included in this calculation.

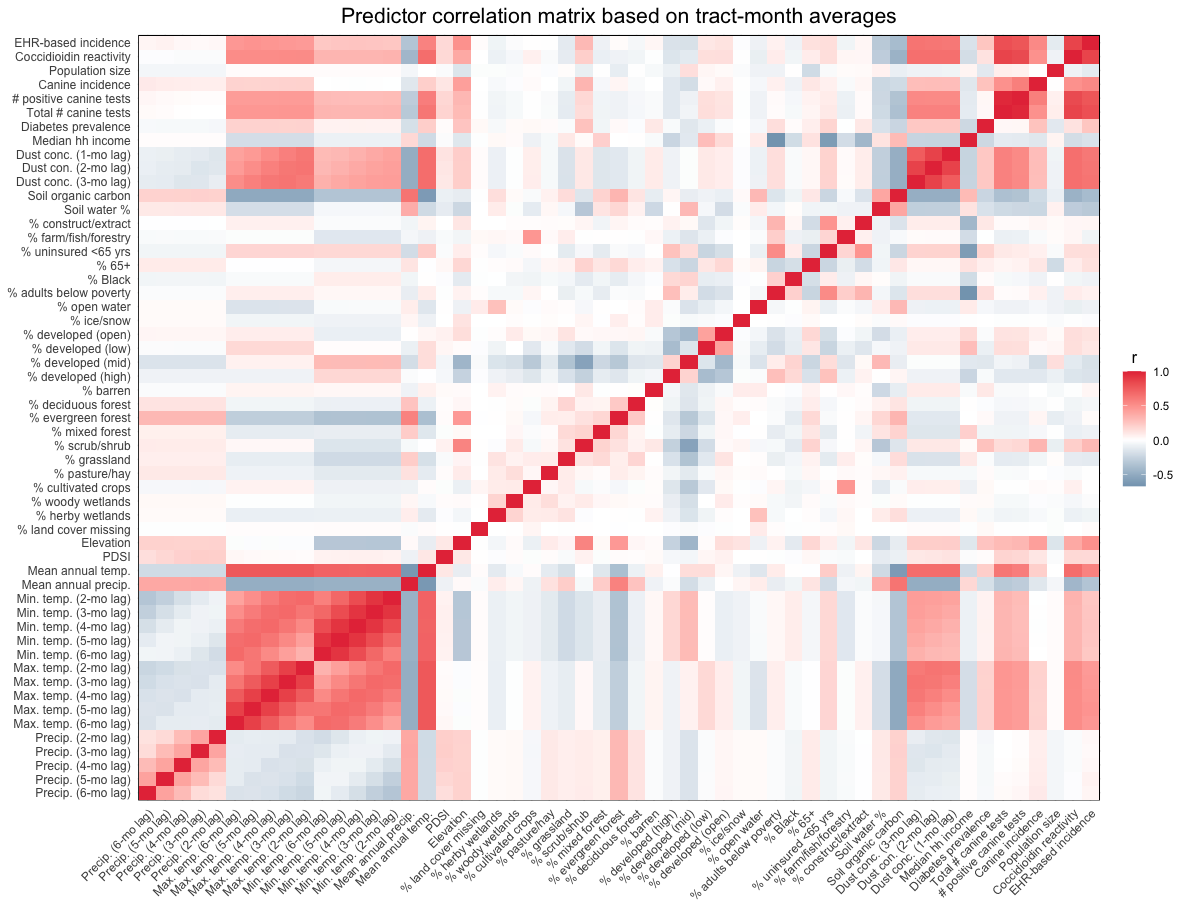

**eFigure 3. Predictor correlation matrix.** Correlations between predictors included in the model based on monthly, tract-level data for Arizona and California. Correlations here were computed as the average across months for a given tract, then averaged across all tracts. Note that we show only the 2- to 6-month lagged temperature and precipitation variables to aid visualization, but the model included 2- to 26-month lags.

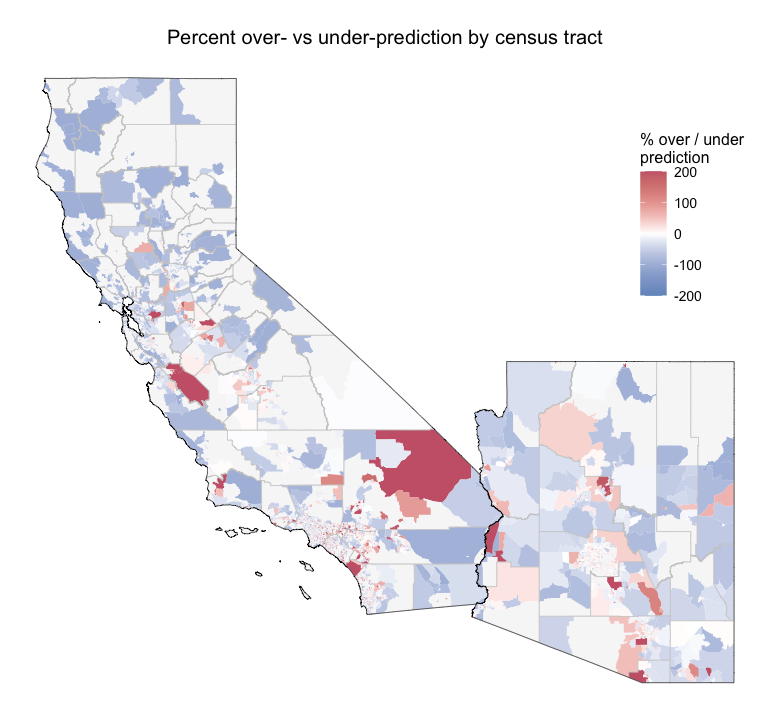

**eFigure 4**. **Spatial bias by census tract**. Spatial bias in model predictions (out-of-sample predictions averaged across 100 bootstrap iterations), expressed as the extent of over- or underprediction in predicted coccidioidomycosis cases relative to reported cases captured in state surveillance. Here, cases are totaled across all months from 2013–2023. Gray denotes census tracts either excluded from the analysis due to small population sizes (<100 people), or for which bias could not be calculated due to zero reported cases across the study period.

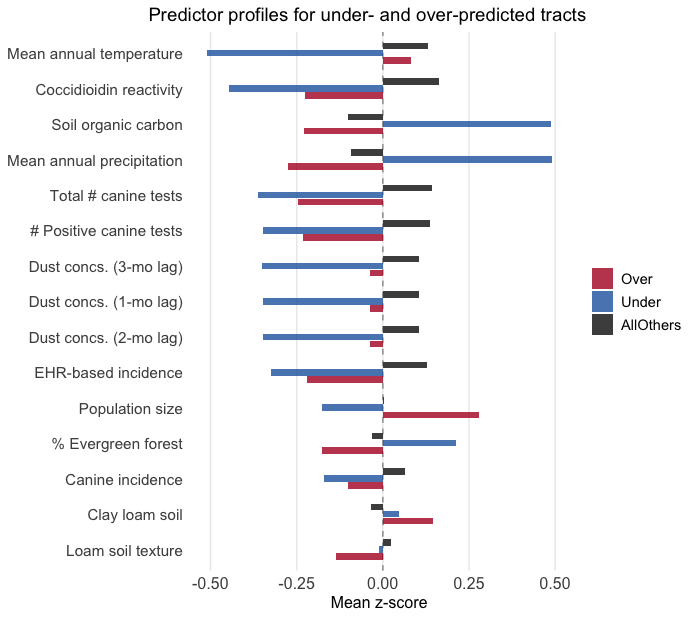

**eFigure 5.** **Features of over- or underpredicted census tracts.** Features of census tracts wherein estimated coccidioidomycosis cases are over- or underpredicted relative to reported cases from state surveillance. Here, the extent of over- or underprediction is based on total cases across all months from 2013–2023. “Overpredicted” and “underpredicted” tracts are those wherein predicted cases are at least 50% higher or lower than reported cases, respectively. All other tracts (“AllOthers”) are included as a reference group. Bars show mean standardized predictor values across tract-months, with positive values indicating that a group’s mean is above the overall mean, and negative values indicating the opposite. Features are ordered on the y-axis based on their absolute standardized difference between over- or underpredicted tracts and the reference, with only the top 15 such features shown.

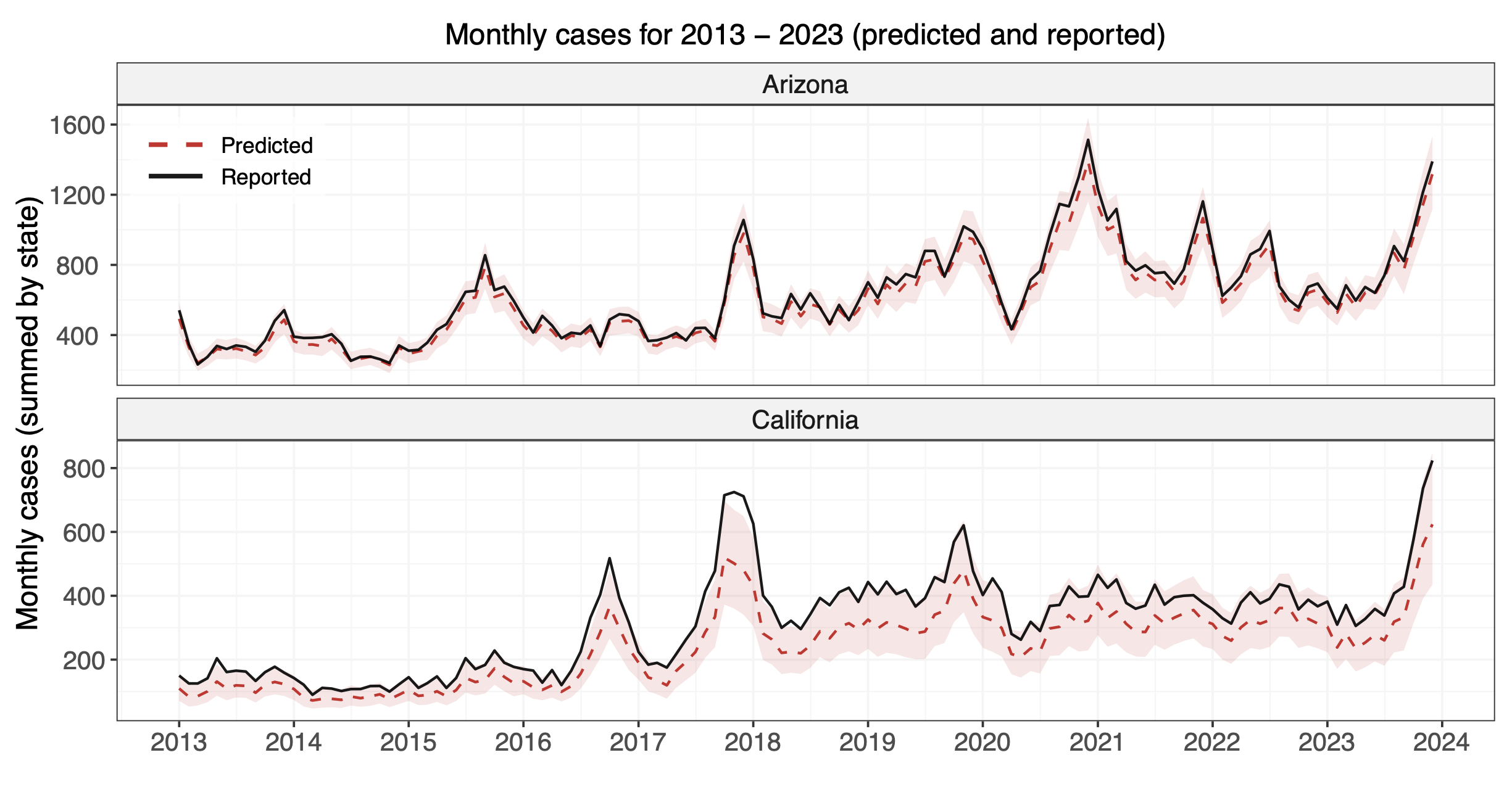

**eFigure 6. Predicted and reported monthly case counts.** Time series of monthly coccidioidomycosis cases from January 2013 to December 2023 for Arizona (top) and California (bottom) based on model predictions (red) and reported cases (black). The dashed red line and light red shading denote the mean and the 95% bootstrap interval, respectively, of the model’s out-of-sample predictions across 100 bootstrap iterations.

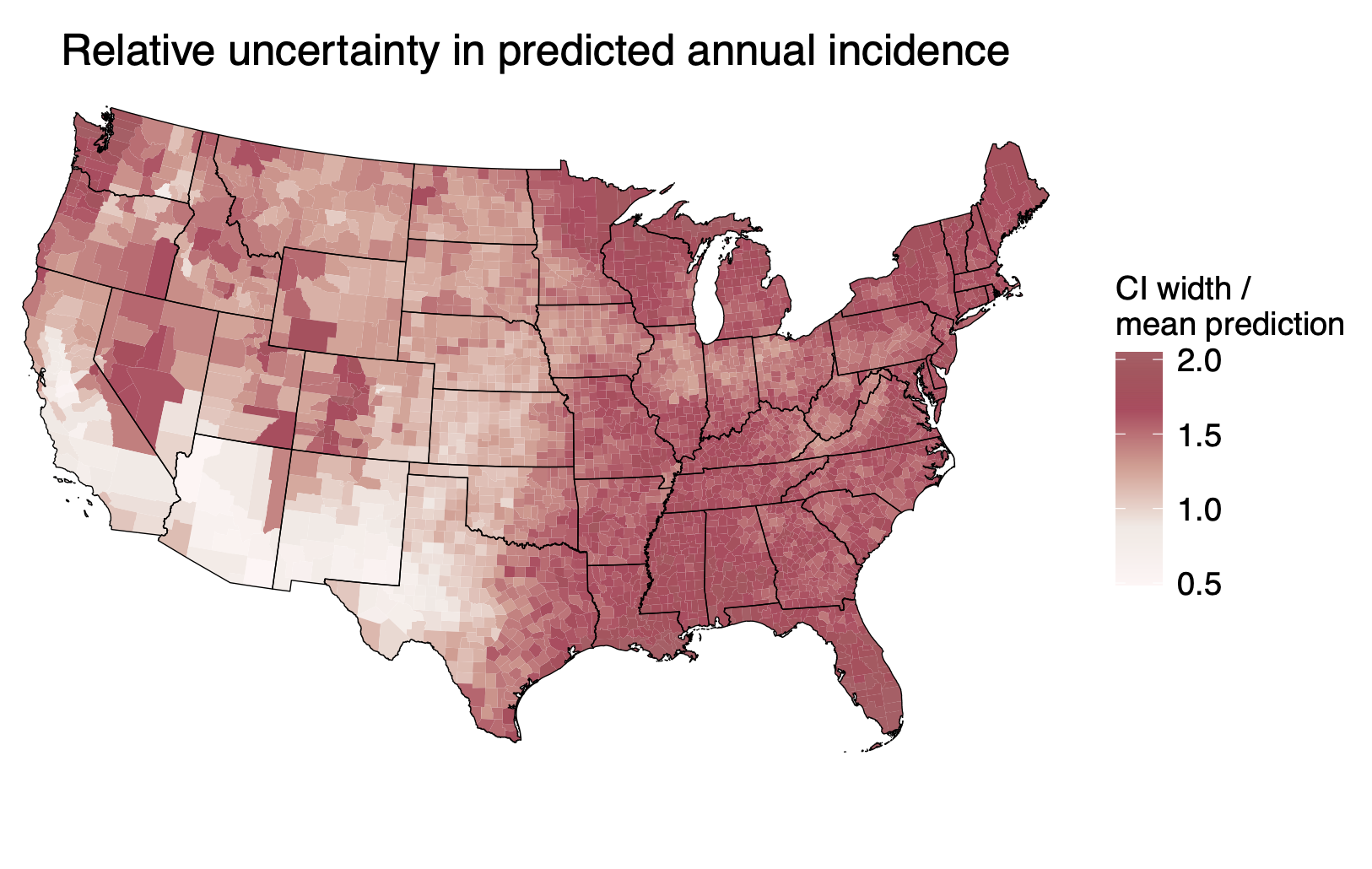

**eFigure 7. Relative uncertainty in predicted annual incidence.** Relative uncertainty is defined here as the width of the 95% bootstrap interval divided by the mean prediction. It is shown here based on predicted county-level mean annual incidence across 2013–2023.

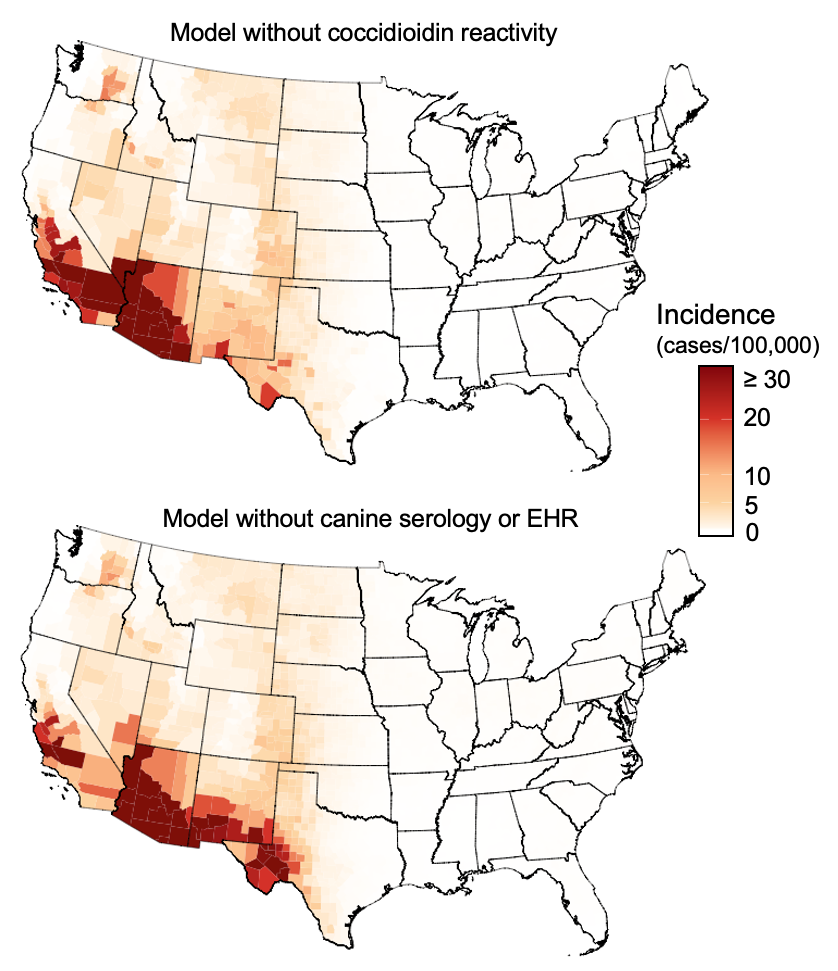

**eFigure 8. Nationwide incidence predictions under alternative model specifications.** Mean annual incidence (cases per 100,000 population) across all US counties as predicted under the alternative model specifications that either excluded coccidioidin reactivity (top) or excluded the canine serology and EHR-based incidence predictors (bottom). Values denote averages across 2013–2023. To aid visualization, incidence values are capped at 30.

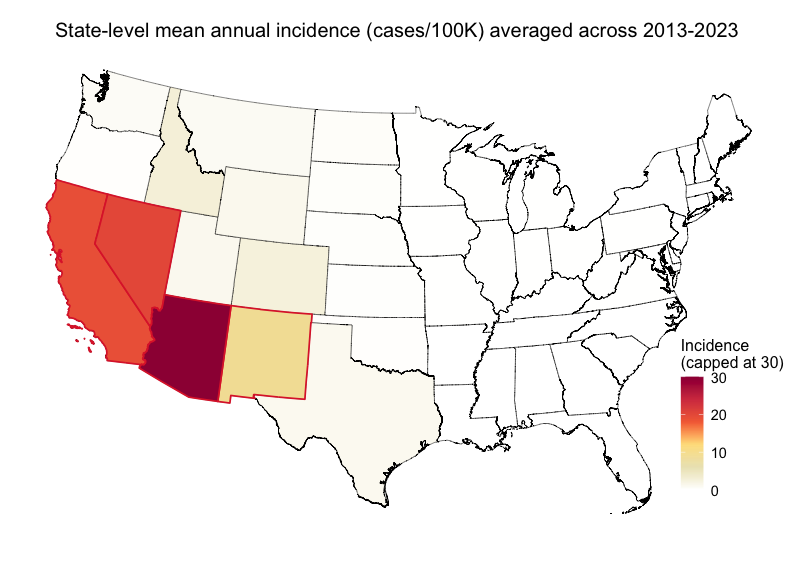

**eFigure 9.** **Predicted state-level mean annual incidence** (cases per 100,000 population). Values denote averages across 2013–2023. To aid visualization, incidence values are capped at 30. States outlined in red (Arizona, California, Nevada, and New Mexico) are those with annual incidence ≥10 cases per 100,000 population for a period of three or more consecutive years—the CDC’s definition of a “high-incidence jurisdiction”^7^.

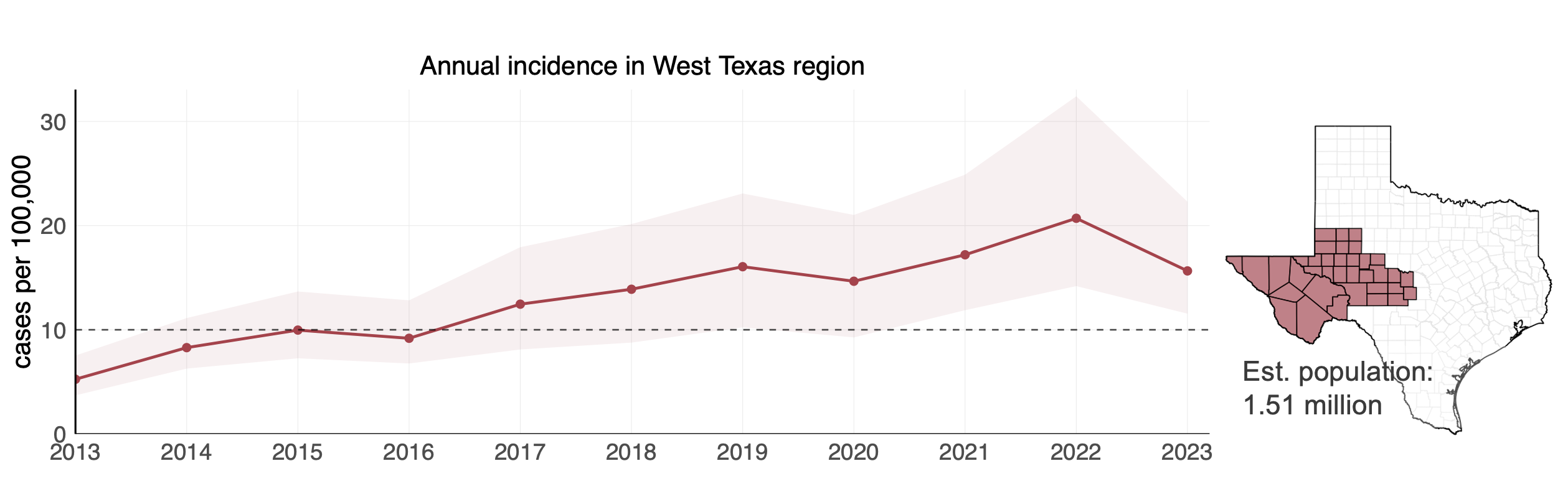

**eFigure 10.** **Mean annual incidence in western Texas.** Time series of predicted annual incidence (cases per 100,000 population) for western Texas—the 36-county region^30^ shown on the right, which has an estimated population size of approximately 1.51 million people. The CDC defines “high-incidence jurisdictions” as those with annual incidence ≥10 cases per 100,000 population (denoted by the dashed horizontal line) for a period of three or more consecutive years^7^.

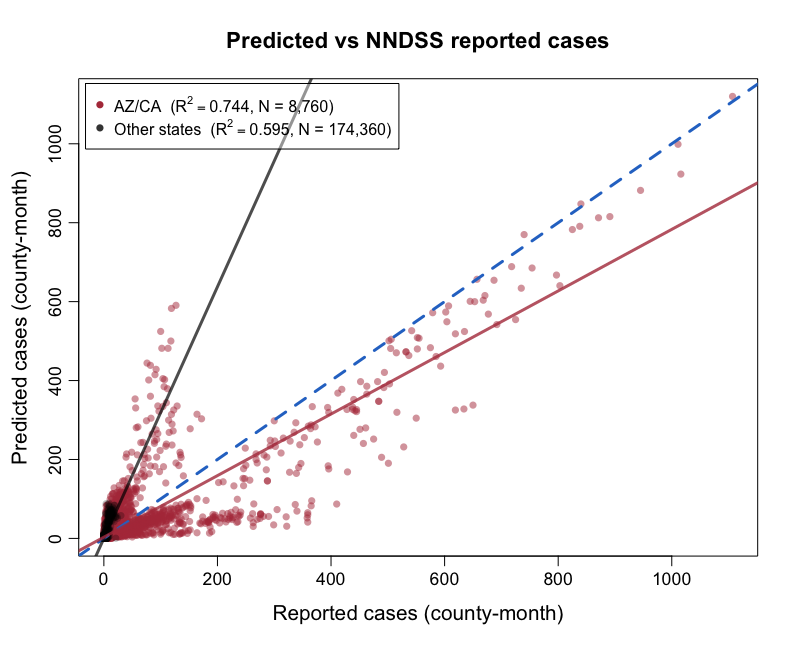

**eFigure 11. Predicted versus NNDSS reported cases.** County-level coccidioidomycosis cases as predicted by the model versus reported to the CDC’s NNDSS. The NNDSS dataset contains case counts for the 28 states with mandatory reporting for coccidioidomycosis. Each point denotes a single county-month between 2013–2022 for Arizona and California (red) or all other states (black). Predicted cases here are the mean across bootstrap iterations (b = 100). The solid lines denote the regression of predicted against reported cases for Arizona and California (red) or all other states (black), while the blue dotted line denotes a 1:1 relationship. The inset text reports the $R^{2}$ between predicted and reported cases and the number of observations included in the calculation.

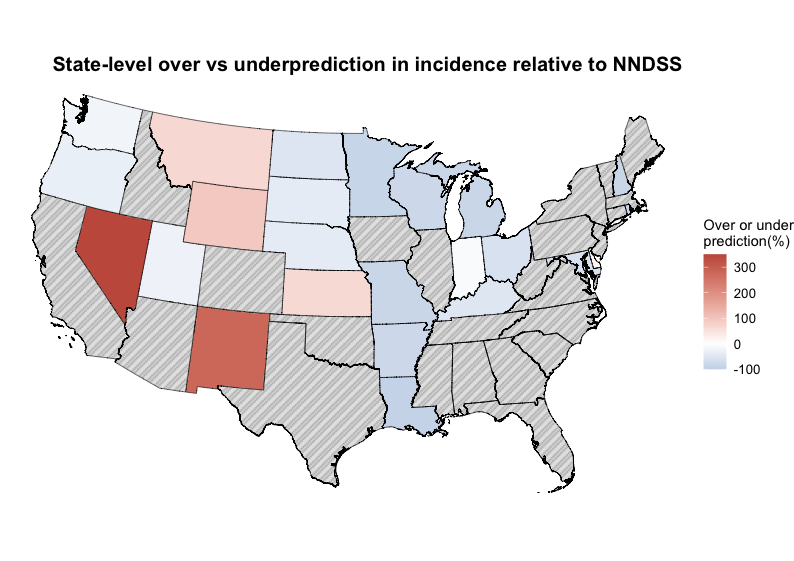

**eFigure 12. Extent of over- or underprediction by state**. Percent difference in state-level annual incidence (averaged across 2013–2022) from model predictions and NNDSS. Overprediction (red) indicates model predictions exceed NNDSS reporting; underprediction (blue) indicates the opposite. States shown in hashed gray are those that were either used in model training and testing (Arizona and California), had zero reported cases between 2013–2022 (Colorado and the District of Columbia), or lack mandatory reporting (all other states).

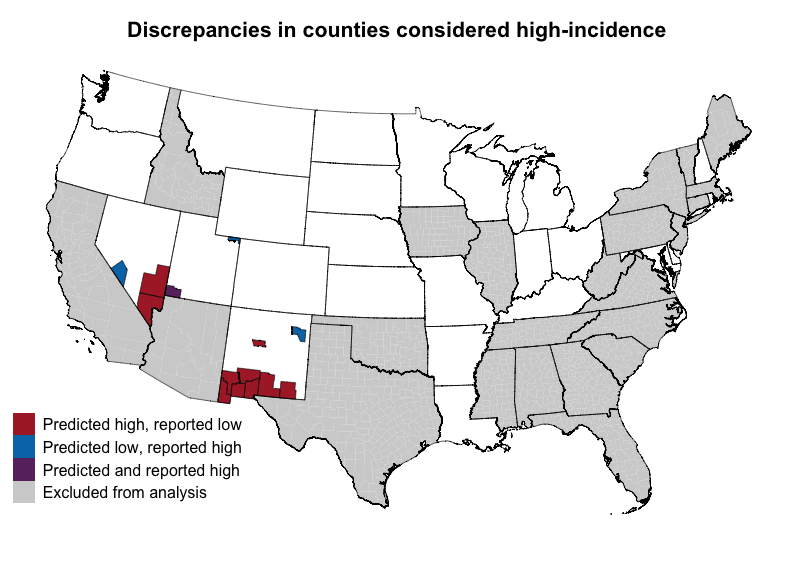

**eFigure 13. Discrepancies in counties considered high-incidence.** Discrepancies between model predictions and NNDSS in which counties are considered “high-incidence” (defined as ≥10 cases per 100,000 population on average across 2013–2022)^31^. Red denotes counties categorized as high-incidence based on model predictions but not based on NNDSS reports (n = 10 counties); blue denotes the opposite (n = 3 counties); and purple denotes counties considered high-incidence by both. States in gray are those excluded from the analysis either because they were used for model training and testing (Arizona and California) or because they lack mandatory reporting (all other states).

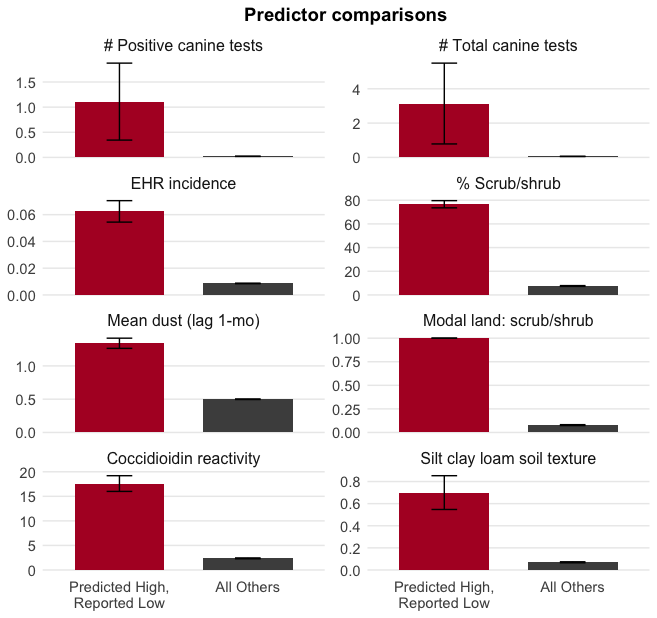

**eFigure 14. Features of counties predicted as high-incidence.** Values of predictors in counties classified as high-incidence based on the mean model predictions but not based on NNDSS case reports (n = 10), and all other counties outside of Arizona and California (n = 3,025). High-incidence was defined as ≥10 cases per 100,000 population on average across 2013–2022. Only the top eight predictors with the largest differences between groups are shown here. The NNDSS dataset contains case counts for the 28 states with mandatory reporting for coccidioidomycosis. Counties in Arizona and California are not included in this feature comparison because these states were used for model training and testing.

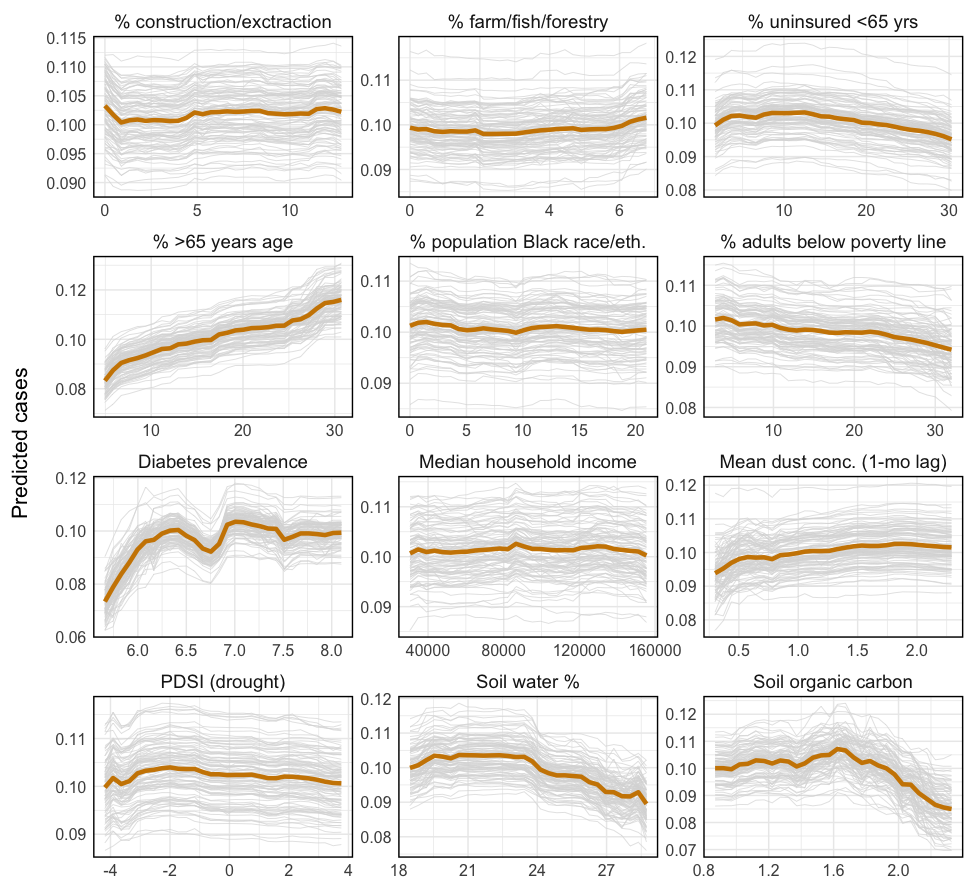

**eFigure 15. Partial dependence plots for additional model predictors.** Partial dependence plots showing the response of predicted cases to a given feature averaged across all possible feature combinations. The orange line denotes the mean response across 100 bootstrap iterations, while the gray lines denote the response from each iteration. Predictors shown here are a subset of those included in the model. See main text Figure 4 for partial dependence plots for additional predictors.

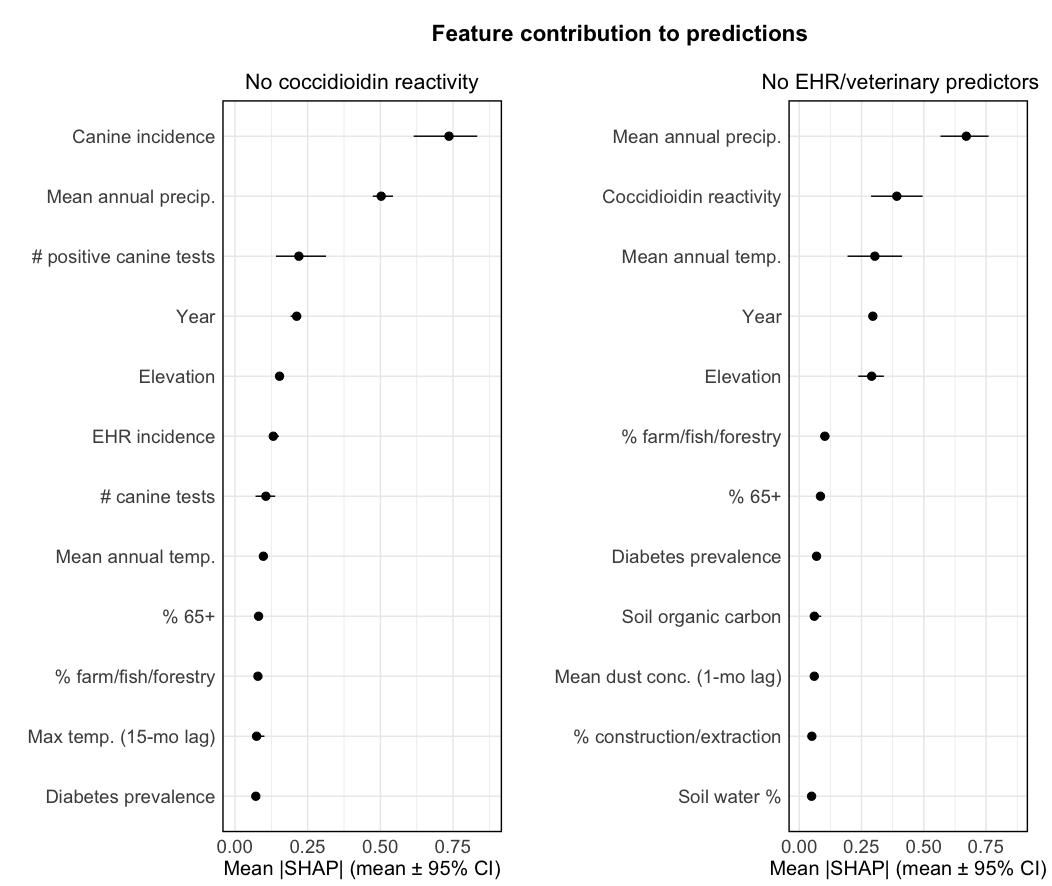

**eFigure 16. Feature importance in alternative model specifications.** Feature contribution to coccidioidomycosis case predictions under the two alternative model specifications that either excluded coccidioidin reactivity as a predictor (left) or excluded the canine serology and EHR-based incidence predictors (right). Points are the mean marginal contribution of a given feature to model predictions across all possible feature combinations (|SHAP| value). Error bars denote the 95% bootstrap interval of marginal contributions computed across 10 bootstrap iterations. Only the top 12 predictors are shown here.

**eTable 1.** **Model hyperparameters tuned using Bayesian optimization.**

| **XGBoost hyperparameter** | **Definition** | **Range of values considered** | **Final value** |
| --- | --- | --- | --- |
| eta | Reduction in influence of each new tree on the existing prediction (also called learning rate) | 0.03–0.15 | 0.03 |
| max_depth | Maximum tree depth | 3–10 | 8 |
| gamma | Minimum loss reduction required to make a tree split | 1–10 | 8.8 |
| min_child_weight | Minimum weight of observations (or minimum number of samples in unweighted data) required to make a tree split | 2–20 | 17 |
| subsample | Proportion of the training data randomly sampled when building each new tree | 0.5–1.0 | 0.63 |
| colsample_bytree | Proportion of available features randomly sampled when building each new tree | 0.5–1.0 | 0.69 |

**eTable 2. Additional model performance metrics**. Performance metrics pertain to the main model specification. Metrics are calculated using out-of-sample predictions (for each tract, the mean across 100 bootstrap iterations in which it was withheld) for Arizona and California at various temporal and spatial aggregations (tract or county, month or year).

| **Level** | **n** | $\boldsymbol{R}^{\boldsymbol{2}}$ | **RMSE** | **Tweedie deviance** |
| --- | --- | --- | --- | --- |
| Tract-month | 1,132,537 | 0.291 | 0.363 | 0.663 |
| Tract-year | 94,391 | 0.681 | 1.806 | 1.364 |
| County-month | 9,636 | 0.996 | 5.243 | 1.113 |
| County-year | 803 | 0.998 | 50.840 | 3.382 |

**eTable 3.** **Performance metrics under various model specifications.** Metrics pertain to: the main model; a model without the coccidioidin reactivity predictor; and a model without the EHR-based incidence or veterinary predictors (i.e., canine incidence, number of positive canine serological tests and total number of canine serological tests). Model performance was evaluated based on the withheld, testing set from Arizona and California at the tract- or county-month level. The top five most important features are determined based on the mean |SHAP| values.

| **Model** | **Tract-month** $\boldsymbol{R}^{\boldsymbol{2}}$ | **Tract-month RMSE** | **County-month** $\boldsymbol{R}^{\boldsymbol{2}}$ | **County-month RMSE** | **Over / under prediction** | **Top five most important features** |
| --- | --- | --- | --- | --- | --- | --- |
| Main | 0.291 | 0.363 | 0.996 | 5.243 | 11.9% under | canine incidence, mean annual precip., coccidioidin reactivity, # positive canine tests, elevation |
| No coccidioidin reactivity | 0.289 | 0.375 | 0.992 | 4.231 | 12.5% under | canine incidence, mean annual precip., # positive canine tests, elevation, EHR-based incidence |
| No EHR or canine serology | 0.285 | 0.377 | 0.991 | 4.573 | 13.8% under | mean annual precip., coccidioidin reactivity, mean annual temp., elevation, % farming/fishing/forestry |

**eTable 4.** **Incidence predictions under various model specifications.** Predictions are shown for the main model and the two alternative specifications that either excluded coccidioidin reactivity as a predictor or excluded the canine serology and EHR-based incidence predictors. Counties are classified as “high incidence” or “moderate incidence” if they have ≥10 or 2–10 cases per 100,000 population^31^, respectively, on average across the period 2013–2023. We list the number of counties within each state (denoted by two-letter state abbreviation) classified into these incidence categories. High-incidence states are those with ≥10 cases per 100,000 population for a period of three or more consecutive years^7^.

|  | **Main** | **No coccidioidin reactivity** | **No EHR or canine serology** |
| --- | --- | --- | --- |
| # of high- incidence counties | AZ (14), CA (17), NV (2), NM (8), TX (8), UT (1) | AZ (14), CA (21), CO (1), ID (1), NV (1), NM (6), OR (1), TX (7), WA (4) | AZ (14), CA (11), NV (1), NM (12), TX (21), UT (1), WA (3) |
| # of moderate- incidence counties | AZ (1), CA (11), CO (20), ID (9), KS (2), MT (11), NE (3), NV (7), NM (21), OK (1), OR (4), TX (42), UT (7), WA (7), WY (5) | AZ (1), CA (6), CO (26), ID (9), KS (6), MT (21), NE (6), NV (8), NM (24), OK (2), OR (3), SD (1), TX (45), UT (12), WA (3), WY (9) | AZ (1), CA (17), CO (24), ID (12), KS (21), MT (19), NE (16), NV (10), NM (18), ND (2), OK (3), OR (4), SD (9), TX (57), UT (10), WA (7), WY (8) |
| high-incidence states | AZ, CA, NV, NM | AZ, CA, NV, NM | AZ, CA, NV, NM |

**eTable 5.** **State-level incidence and cases from model predictions and NNDSS.** Incidence is expressed as cases per 100,000 population. For the predicted case and incidence values, we report the mean and 95% bootstrap interval (in brackets). Values denote averages across 2013–2022. We also report the extent of under- or overprediction in incidence for each state, calculated based on annual state-level averages across this time period (positive values indicate predictions exceed reports; negative values indicate the opposite). Arizona and California are excluded from this comparison as these states were used for model training and testing. For Colorado and the District of Columbia, prediction discrepancies could not be calculated as no cases were reported there between 2013–2022. The CDC’s NNDSS dataset contains data for the 28 states with mandatory reporting for coccidioidomycosis; states without mandatory reporting are denoted with (*).

|  | **Mean annual cases** | | **Mean annual incidence** | | **Under- or overprediction (%)** |
| --- | --- | --- | --- | --- | --- |
| **State** | Predicted | NNDSS | Predicted | NNDSS |  |
| Alabama* | 0.8 [0.3, 1.2] | NA | 0.02 [0.01, 0.02] | NA | NA |
| Arizona | 6184.2 [4543.2, 7498.6] | 8246.1 | 87.94 [64.60, 106.50] | 117.94 | NA |
| Arkansas | 0.8 [0.4, 1.2] | 4.3 | 0.03 [0.01, 0.04] | 0.14 | -81.6 |
| California | 7590.0 [5764.1, 9621.8] | 5927.4 | 19.33 [14.68, 24.51] | 15.1 | NA |
| Colorado | 137.6 [92.8, 191.3] | 0 | 2.42 [1.63, 3.36] | 0 | — |
| Connecticut* | 0.8 [0.4, 1.3] | NA | 0.02 [0.01, 0.04] | NA | NA |
| Delaware | 0.2 [0.1, 0.4] | 0.2 | 0.02 [0.01, 0.04] | 0.02 | 15.8 |
| District of Columbia | 0.3 [0.2, 0.6] | 0 | 0.05 [0.02, 0.08] | 0 | — |
| Florida* | 3.2 [1.6, 5.4] | NA | 0.02 [0.01, 0.03] | NA | NA |
| Georgia* | 3.0 [1.4, 5.0] | NA | 0.03 [0.01, 0.05] | NA | NA |
| Idaho* | 48.8 [29.9, 69.2] | NA | 2.73 [1.67, 3.86] | NA | NA |
| Illinois* | 9.8 [5.7, 16.2] | NA | 0.08 [0.04, 0.13] | NA | NA |
| Indiana | 3.5 [1.8, 5.9] | 4 | 0.05 [0.03, 0.09] | 0.06 | -11.5 |
| Iowa* | 2.5 [1.4, 3.8] | NA | 0.08 [0.05, 0.12] | NA | NA |
| Kansas | 5.4 [3.9, 7.5] | 3.3 | 0.18 [0.13, 0.25] | 0.11 | 63.2 |
| Kentucky | 1.5 [0.7, 2.6] | 3.2 | 0.03 [0.02, 0.06] | 0.07 | -52.0 |
| Louisiana | 0.5 [0.3, 0.8] | 10 | 0.01 [0.01, 0.02] | 0.21 | -95.1 |
| Maine* | 0.2 [0.1, 0.4] | NA | 0.02 [0.01, 0.03] | NA | NA |
| Maryland | 2.1 [1.1, 3.8] | 5.6 | 0.04 [0.02, 0.06] | 0.09 | -61.9 |
| Massachusetts* | 1.4 [0.7, 2.2] | NA | 0.02 [0.01, 0.03] | NA | NA |
| Michigan | 5.0 [2.7, 8.4] | 33.3 | 0.05 [0.03, 0.08] | 0.33 | -85.0 |
| Minnesota | 3.6 [1.9, 5.3] | 30.5 | 0.06 [0.03, 0.09] | 0.54 | -88.1 |
| Mississippi* | 0.3 [0.2, 0.5] | NA | 0.01 [0.01, 0.02] | NA | NA |
| Missouri | 3.0 [1.4, 4.8] | 21.3 | 0.05 [0.02, 0.08] | 0.35 | -86.0 |
| Montana | 8.3 [5.2, 12.1] | 4.8 | 0.77 [0.48, 1.13] | 0.46 | 67.9 |
| Nebraska | 5.2 [3.8, 6.8] | 8.7 | 0.27 [0.20, 0.35] | 0.45 | -40.3 |
| Nevada | 617.5 [371.6, 915.7] | 136.6 | 20.41 [12.29, 30.19] | 4.54 | 349.6 |
| New Hampshire | 0.3 [0.1, 0.5] | 2.1 | 0.02 [0.01, 0.03] | 0.15 | -87.2 |
| New Jersey* | 2.3 [1.3, 3.8] | NA | 0.03 [0.01, 0.04] | NA | NA |
| New Mexico | 185.1 [145.5, 232.8] | 48.7 | 8.78 [6.90, 11.04] | 2.31 | 279.9 |
| New York* | 6.6 [3.8, 11.0] | NA | 0.03 [0.02, 0.05] | NA | NA |
| North Carolina* | 2.9 [1.5, 4.6] | NA | 0.03 [0.01, 0.04] | NA | NA |
| North Dakota | 2.9 [1.8, 4.5] | 6.2 | 0.38 [0.24, 0.58] | 0.81 | -53.6 |
| Ohio | 7.0 [3.7, 11.6] | 18.1 | 0.06 [0.03, 0.10] | 0.15 | -61.3 |
| Oklahoma* | 5.1 [3.3, 7.3] | NA | 0.13 [0.08, 0.19] | NA | NA |
| Oregon | 5.5 [3.6, 8.2] | 8.3 | 0.13 [0.09, 0.20] | 0.2 | -34.5 |
| Pennsylvania* | 5.8 [3.1, 9.6] | NA | 0.04 [0.02, 0.07] | NA | NA |
| Rhode Island | 0.2 [0.1, 0.3] | 1.2 | 0.01 [0.01, 0.03] | 0.11 | -86.6 |
| South Carolina* | 1.3 [0.7, 1.9] | NA | 0.03 [0.01, 0.04] | NA | NA |
| South Dakota | 2.5 [1.8, 3.7] | 4.3 | 0.29 [0.20, 0.42] | 0.49 | -41.2 |
| Tennessee* | 1.7 [0.7, 2.6] | NA | 0.02 [0.01, 0.04] | NA | NA |
| Texas* | 293.6 [221.9, 412.9] | NA | 1.02 [0.77, 1.44] | NA | NA |
| Utah | 38.1 [25.9, 52.8] | 51.4 | 1.19 [0.81, 1.64] | 1.63 | -27.3 |
| Vermont* | 0.1 [0.1, 0.2] | NA | 0.02 [0.01, 0.03] | NA | NA |
| Virginia* | 2.9 [1.6, 4.5] | NA | 0.03 [0.02, 0.05] | NA | NA |
| Washington | 42.2 [25.7, 71.2] | 54.8 | 0.56 [0.34, 0.94] | 0.72 | -22.2 |
| West Virginia* | 1.1 [0.6, 1.7] | NA | 0.06 [0.03, 0.09] | NA | NA |
| Wisconsin | 2.7 [1.5, 4.5] | 14.7 | 0.05 [0.03, 0.08] | 0.25 | -81.4 |
| Wyoming | 6.9 [4.0, 10.5] | 3.6 | 1.20 [0.69, 1.81] | 0.62 | 93.1 |

11.	Centers for Disease Control and Prevention. Diabetes Data and Statistics. Published online 2024.

12.	Arizona Office of Economic Opportunity. Population Estimates. Published online 2025.

13.	State of California Department of Finance. Population and Housing Estimates. Published online 2025.

14.	Weaver AK, Keeney N, Head JR, et al. Estimating the Exposure–Response Relationship between Fine Mineral Dust Concentration and Coccidioidomycosis Incidence Using Speciated Particulate Matter Data: A Longitudinal Surveillance Study. *Environ Health Perspect*. 2025;133(1):017003. doi:10.1289/EHP13875

15.	Van Donkelaar A, Martin RV, Li C, Burnett RT. Regional Estimates of Chemical Composition of Fine Particulate Matter Using a Combined Geoscience-Statistical Method with Information from Satellites, Models, and Monitors. *Environ Sci Technol*. 2019;53(5):2595-2611. doi:10.1021/acs.est.8b06392

16.	Hengl T, Consoli D, Tian X, et al. OpenLandMap-soildb: global soil information at 30 m spatial resolution for 2000–2022+ based on spatiotemporal Machine Learning and harmonized legacy soil samples and observations. *ESSD – Land/Pedology*. Preprint posted online June 24, 2025. doi:10.5194/essd-2025-336

17.	United States Department of Agriculture. Soil Taxonomy A Basic System of Soil Classification for Making and Interpreting Soil Surveys. Published online 1999.

18.	Dewitz J. US Geological Survey, 2021, National Land Cover Database (NLCD) 2019 Products (ver. 2.0, June 2021): US Geological Survey data release. Published online 2023.

19.	United States Geological Survey. USGS EROS Archive - Digital Elevation - Global Multi-resolution Terrain Elevation Data 2010. Published online 2018.

20.	PRISM Group, Oregon State University. PRISM Weather Data. Published online 2014. https://prism.oregonstate.edu

21.	Daly C, Halbleib M, Smith JI, et al. Physiographically sensitive mapping of climatological temperature and precipitation across the conterminous United States. *International Journal of Climatology*. 2008;28(15):2031-2064. doi:10.1002/joc.1688

22.	Head JR, Sondermeyer-Cooksey G, Heaney AK, et al. Effects of precipitation, heat, and drought on incidence and expansion of coccidioidomycosis in western USA: a longitudinal surveillance study. *The Lancet Planetary Health*. 2022;6(10):e793-e803. doi:10.1016/S2542-5196(22)00202-9

23.	National Oceanic and Atmospheric Administration, University of California, Merced. Gridded Surface Meteorological (gridMET) Dataset.

24.	Chen T, Guestrin C. XGBoost: A Scalable Tree Boosting System. Published online August 13, 2016. Accessed June 1, 2022. https://dl.acm.org/doi/10.1145/2939672.2939785

25.	Vihta KD, Pritchard E, Pouwels KB, et al. Predicting future hospital antimicrobial resistance prevalence using machine learning. *Commun Med*. 2024;4(1):197. doi:10.1038/s43856-024-00606-8

26.	Bennett J, Lanning S. The Netflix Prize. In: *Proceedings of KDD Cup and Workshop*. 2007.

27.	Rusdah DA, Murfi H. XGBoost in handling missing values for life insurance risk prediction. *SN Applied Sciences*. 2020;2.

28.	Kokonendji CC, Bonat WH, Abid R. Tweedie regression models and its geometric sums for (semi‐)continuous data. *WIREs Computational Stats*. 2021;13(1):e1496. doi:10.1002/wics.1496

29.	Yan Y. rBayesianOptimization: Bayesian Optimization of Hyperparameters. Published online 2025.

30.	Jordan TG. Perceptual Regions in Texas. *Geographical Review*. 1978;68(3):293. doi:10.2307/215048

31.	Hurd-Kundeti G, Sondermeyer Cooksey GL, Jain S, Vugia DJ. Valley Fever (Coccidioidomycosis) Awareness — California, 2016–2017. *MMWR Morb Mortal Wkly Rep*. 2020;69(42):1512-1516. doi:10.15585/mmwr.mm6942a2
